# A walk in the park – identifying healthy greenspaces using scents

**DOI:** 10.1101/2025.08.27.25334443

**Authors:** William T. Kay, Anya Lindström Battle, Mahal Humberstone, Molly Tucker, Kieran E. Storer, Geoffrey Kite, Katherine Willis

**Affiliations:** Department of Biology, University of Oxford, South Parks Road, Oxford OX1 3RB, UK; Jodrell Laboratory, Royal Botanic Gardens, Kew, Richmond, Surrey TW9 3AB, UK

**Keywords:** Greenspace chemical ecology, Gas Chromatography-Mass Spectrometry, volatile analysis, Volatile Organic Compounds, Biogenic Volatile Organic Compounds, terpenes, health

## Abstract

1.

1. Biogenic volatile organic compounds (bVOCs) emitted by plants can promote stress reduction and cognitive benefits, whereas anthropogenic VOCs (aVOCs) common in urban air, such as BTEX compounds, pose health risks. We aimed to characterise and compare these airborne chemicals across urban greenspaces and develop a novel, health-oriented metric for site evaluation.
2. Air samples were collected across six Oxford greenspaces on a single date using Tenax™ filters and analysed by GC–MS. Data were processed using metabolomics-style workflows, identifying 245 biologically relevant compounds. Multivariate analyses compared site profiles, and one site was monitored over 12 months to assess environmental influences.
3. Greenspaces exhibited significantly different volatile signatures, including variation in both health-promoting bVOCs and harmful aVOCs. Temperature, humidity, wind speed, and rainfall significantly influenced the presence and abundance of beneficial compounds.
4. These results highlight the importance of considering airborne chemistry in urban planning and public health decision-making. Integrating VOC profiling into planning and public health strategies could support delivery of higher-quality, health-promoting urban environments and inform guidance on optimal timing for outdoor activity.

**Societal Impact Statement:** As urbanisation accelerates globally, access to nature is increasingly recognised as vital for public health and wellbeing. We captured and analysed plant-emitted airborne “scent signatures” across Oxford’s urban greenspaces to assess their potential health relevance. We found that sites differ in levels of health-promoting volatiles and harmful pollutants, and that these compounds vary with environmental conditions such as temperature and humidity. Our findings provide a novel framework for evaluating urban greenspaces, informing planning, air-quality management, and public health strategies to create greener, healthier cities.

## 3. Introduction

Biogenic volatile organic compounds (bVOCs) are a chemically diverse group of carbon-based secondary metabolites produced across all kingdoms of life (Wang et al. 2024). They function in stress protection, signalling, and communication within and between species (Weisskopf, Schulz, and Garbeva 2021) and many are released into the air via passive diffusion from different tissues. In plants, bVOCs such as isoprene, monoterpenes, and sesquiterpenes account for almost all biogenic emissions (Guenther et al. 2012). Among these, monoterpenes such as alpha-pinene, beta-pinene, and limonene are key contributors to both atmospheric chemistry and human well-being. Numerous studies link exposure to plant-emitted terpenes with lowered cortisol, blood pressure, and heart rate, as well as improved mood, cognition, and immune function (Zeng et al. 2022; Park et al. 2010; Li 2010; Bratman et al. 2015). By contrast, anthropogenic VOCs (aVOCs) derived from transport, plastics, and industrial sources - particularly benzene, toluene, ethylbenzene, and xylene (collectively known as the BTEX compounds) - are well-known urban pollutants associated with respiratory and cardiovascular diseases and cancer (Gao et al. 2023).

Because of their location the ‘scentscapes’ (the total volatile profile in a specific time and place) of urban greenspaces are likely to comprise both bVOCs and aVOCs. Understanding the balance between these “healthy” and “unhealthy” volatiles is therefore essential for assessing how urban greenspaces influence well-being, and for driving policy for positive changes in urban development.

Here, we present a new analytical pipeline for characterising ambient air VOCs *in situ*, integrating field sampling and thermal desorption gas chromatography mass spectrometry (TD-GC-MS) analysis. Although similar methods have been developed for headspace sampling (Yin et al. 2022; de Carvalho Couto et al. 2024; Reyrolle et al. 2024), none have yet been published for the analysis of ambient air. We apply this approach to six Oxford greenspaces to test whether site location determines the distribution of biogenic versus anthropogenic volatiles - and to identify those environments most likely to promote human health.

Additionally, it is well known that the release of bVOCs varies in time. For instance, plants coordinate their volatile production with the availability of substrates from photosynthesis, which varies considerably over the course of a day. Indeed, the production of most plant volatiles is under diurnal and/or circadian control; on a longer timescale, the effects of factors such as seasonality, phenology, and other environmental effects including temperature and humidity on the emission of volatiles have been well established (Joo et al. 2018; Yang et al. 2024; Mu et al. 2022; Li et al. 2023).

Firsly, we analysed volatiles on a single date as a snapshot across six outdoor greenspace sites. Secondly, we sampled a single site – the Botanical Garden outdoors – over a full year, linking differences in volatile fingerprints to seasonal environmental changes and transient weather events. Our work aimed to assess three main hypotheses: (i) that a site’s location is a significant driver of volatile difference enabling the separation of greenspaces by their scentscape, (ii) that urban greenspace volatile profiles change over time dependent on environmental variables, and iii) there is sufficient distinction between the presence and abundance of both bVOCs and aVOCs between urban green spaces that it is possible to determine which may be more healthy/unhealthy for human recreation.

## 4. Materials and Methods

### 4.1 Site locations and sampling strategy

Six urban greenspace sites around Oxford were chosen as sampling sites due to their (i) public accessibility, (ii) different ecologies, and (iii) contrasting distance from the city centre. We selected five outdoor greenspaces (Botanic Garden Outdoors, University Parks Dense, University Parks Open, Warneford Meadow, and Wytham Woods) and one indoor site (the Rainforest Glasshouse at the Oxford Botanical Gardens) (Figure 1A).

**Figure 1:**
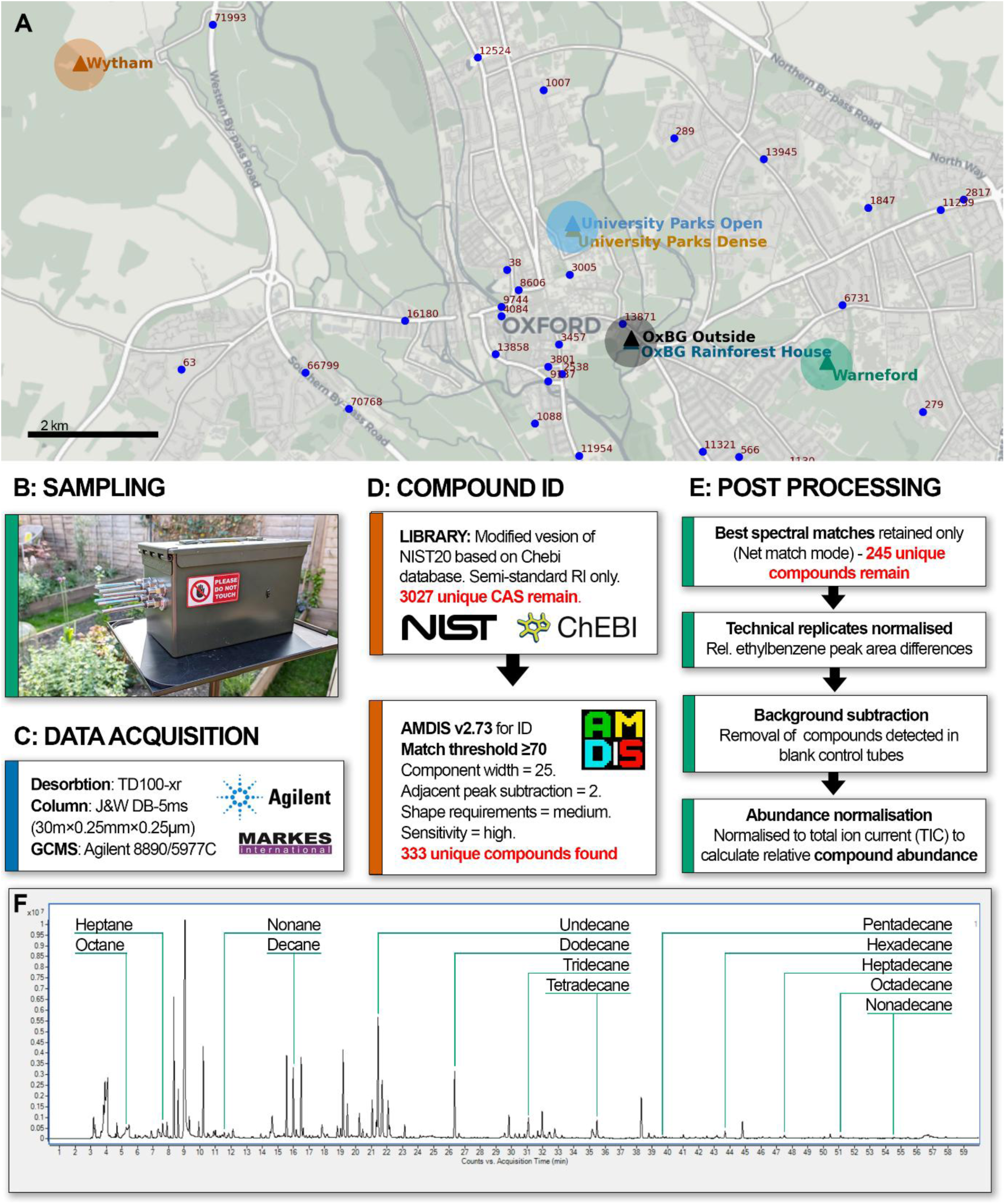
A: Map of the sampling locations used in this study. (Botanic Garden Outdoors; Botanic Garden Rainforest Glasshouse; University Parks Dense; University Parks Open; Warneford Meadow; Wytham Woods). Numbers on roads indicate Annual Average Daily Traffic (AADF) for 2023 as representative data for each site (GOV.UK 2023) B: Sampling box (VOKSBOX) developed for this work on a tripod at 1.5m height. Air from the GilAir® sample pump is split up to 6 ways using brass splitters with silicone tubing. C-E: A flow chart of the methods used in this paper for acquisition, identification, and processing of compounds. Red text shows number of compounds identified after each processing step. F: Example chromatogram resulting from sample collection in Botanical Gardens Outdoors sampling site and GCMS protocol - annotated with positions of alkanes used to build Kovats retention index.

To investigate the characteristic VOC profiles of the selected sites, we employed a collection protocol adapted from Walker et al (2023) with the addition of housing the pump and apparatus within a bespoke metal box (VOKSBOX, B). Ambient air was captured using a GilAir® pump (Sensidyne, LP, St. Petersburg, FL, USA) connected simultaneously to 4 replicate Markes (Markes international Ltd. Bridgend, UK) Tenax® TA tubes using a flow rate of 400ml/min for 2 hours (12 L of air per tube). Samples were collected at a height of 1.5m – based on the approximate level of the adult human nose while walking - between the hours of 11:00 - 13:00 (+/- 1 hour) GMT. Tubes were capped immediately after collection and stored at - 20 °C for up to 14 days before GC-MS analysis.

A pilot dataset was collected on 27 November 2024 and used to assess the hypothesis that sampling sites have distinct volatile profiles (Figure 2). As well as looking at a snapshot of the scentscapes across the six urban greenspaces, we also wanted to explore how these scentscapes change across time and in the face of environmental variation (Figure 3). Because of the known impacts of seasonality on bVOC emissions we focused only on biogenic volatiles. Due to the higher numbers of biogenic volatiles present in the Botanical Outdoors location, and as this was likely to be the site with the highest footfall, we also sampled this site on 20 January 2025, 04 March 2025, 30 April 2025, 30 June 2025, 24 July 2025, and 04 September 2025. This allowed us to capture a range of temperature, humidity, precipitation, and wind conditions (Figure 3A). All Botanical Outdoors samples during the year were taken from 11:00 to 13:00 in exactly the same GPS location and height.

**Figure 2:**
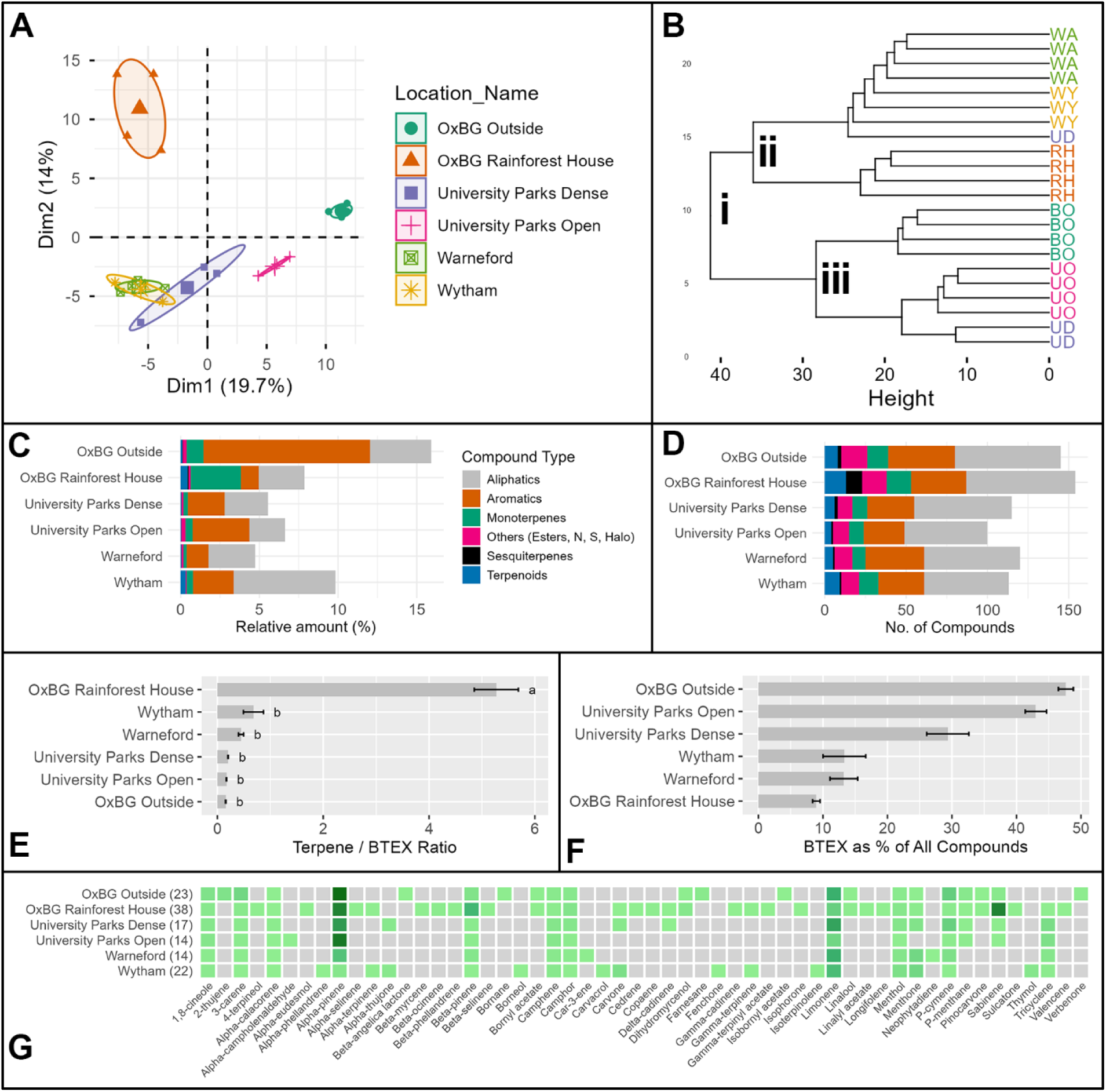
A: PCA which used all 245 identified compounds as predictor variables revealed significant variation between sites, each showing distinctive site-specific clustering (PERMANOVA, F5,16 = 6.557, p = 0.001). B: HCA conducted using Ward’s linkage and Euclidean distances using all technical replicate samples over 6 urban greenspaces. C: Relative abundance (means) of detected volatile compounds split by classes across sites, expressed as a percentage of the total sample. D: Total number of detected volatile compounds per site, categorized by compound class. Results show that the number of all compounds groups differed between sites. E: Terpene: BTEX ratio of sampling sites (∑terpenes/∑BTEX). Relative amounts of all terpenes/terpenoids as a ratio to the anthropogenic BTEX compound. F: ∑BTEX as % of all identified compounds between sampling sites. G: Terpenes/terpenoid compounds identified across sampling locations. Heatmap showing the scaled relative amount (light green > dark green) or absence (grey) of any terpenes/terpenoid VOCs detected across six sampling locations. Scaling was carried out within single locations. C, D, F: Bars represent the mean (n = 4 technical replicate sampling tubes per site). Letters indicate statistically distinct sampling locations within chemical classes based on one-way ANOVA followed by Tukey’s HSD post-hoc test (p < 0.05).

**Figure 3:**
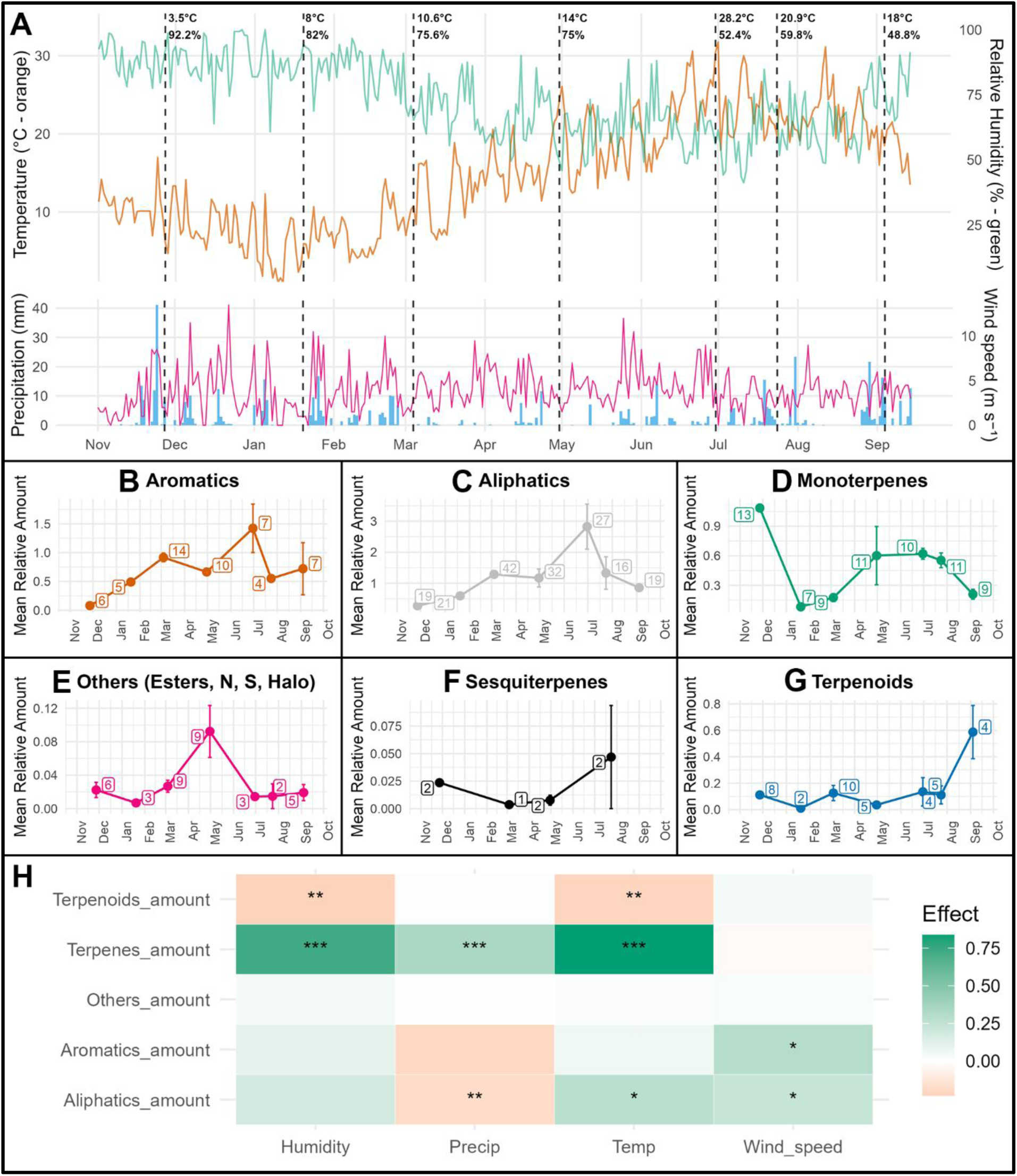
Seasonal dynamics and environmental drivers of biogenic volatile organic compounds (bVOCs) at the Botanical Garden outdoor site. (A) Daily temperature (orange) and relative humidity (green), and daily precipitation (bars) with wind speed (line), derived from local and regional meteorological datasets. Vertical dashed lines indicate sampling dates; values above denote site-specific temperature and relative humidity measured during sampling. (B-G) shows monthly mean relative abundances (± SE) of compound classes (aliphatics, aromatics, monoterpenes, sesquiterpenes, terpenoids, and “others” including esters and N-, S- and halogen-containing compounds) from November 2024 to September 2025. Numbers beside points indicate the number of detected compounds contributing to each class per sampling event. (H) Mixed-effects model coefficients linking standardised environmental predictors (temperature, precipitation, humidity and wind speed) to log-transformed relative abundances of each compound class. Tile colour represents effect size (positive = green, negative = brown), and symbols denote statistical significance (* p < 0.05; ** p < 0.01; *** p < 0.001). Models were fitted with sampling date as a random effect to account for temporal structure.

Site-specific temperature and relative humidity at the time of sampling were recorded using a Kestrel Drop D2 data logger (Nielsen-Kellerman Company, Boothwyn, PA, USA). Historical meteorological data were obtained from multiple sources: air temperature data were retrieved from the Oxford Radcliffe Weather Station, and mean values recorded between 11:00 and 13:00 were used to coincide with the sampling window. Precipitation, humidity, wind speed, and wind direction data were obtained from the Kidlington weather station via Weather Underground (www.wunderground.com).

### 4.2 Identification and analysis of VOC profiles

#### GC-MS Protocol

For identification of VOC profiles (Figure 1C-E), we employed gas chromatography mass spectrometry (GC-MS) analysis at the Jodrell laboratory, Royal Botanic Gardens, Kew. Samples were stored at room temperature for at least 2 hours before desorption using a TD100-xr automated thermal desorption (TD) unit from Markes (Markes international Ltd. Bridgend, UK). Samples were desorbed on to a Tenax® TA packed cold trap (set to 4 °C) by heating the sampling tubes for 10 mins at 230 °C using a flow rate of 50 ml/min helium. Samples were purged from the cold trap onto a J&W DB-5ms GC column (30 m × 0.25 mm × 0.25 µm) within an Agilent 8890 GC System (Agilent Technologies, Santa Clara, CA, USA) by heating from 4 °C to 300 °C over 5 mins at a purge flow rate of 1ml/min Helium. Chromatography was achieved by heating the column at an initial temperature of 40 °C at a rate of 3 °C/min to 220 °C at a flow rate of 1 ml/min Helium. Data were collected in electron ionization (EI) mode using an Agilent 5977C GC/ MSD over a mass range of 50–400 amu.

#### Library Development

The GC-MS library used for identification of compounds was a modified version of the NIST20 Wiley Registry 12th edition mass spectral library minimised to compounds existing in the Chemical Entities of Biological Interest (ChEBI) database (Degtyarenko et al. 2007). Only compounds with entries for retention indexes (RI) on semi-standard non-polar columns (to match the J&W DB-5ms column used) were selected. To avoid mismatches, isomers with the same RI were combined (e.g. (−)-alpha-pinene / (+)-alpha-pinene, became alpha-pinene). The final library used (W12N20MAIN_chebi_WK_edit.MSP) contained 3,027 unique compounds.

#### Pre-Analysis

To aid in the correct identification of compounds, a non-isothermal Kovats index (Kovats 1958) was built using retention times from C8 – C20 alkane standards (Merck KGaA, Darmstadt, Germany,, 04070-1ML, Figure 1F). This index uses known compounds to produce accurate estimations of where all compounds will elute from the GC column (retention times) thus allowing precise identification. We validated this index using 60 pure terpene standards (Restek Corporation, Bellefonte, PA, USA, 34142 and 34143).

#### Compound Identification

Sample analysis was performed using the Automated Mass Spectral Deconvolution and Identification System (AMDIS) version 2.73. Compound identification relied on spectral and RI matching, with a minimum match factor threshold of 70 (Net), a RI window of +/- 10, and a very strong match factor penalty. Deconvolution settings were component width: 25; adjacent peak subtraction: two; shape requirements: medium; sensitivity: high; and resolution: medium. The analysis was restricted to a single identification per library compound.

#### Post Processing

The relative amount (%) of each compound within a sample was used for all analysis in this paper. This was calculated as based on the Total Ion Count (TIC) of the sample / peak area of each compound. No absolute quantification was carried out in this work due to not having concentration curves for all compounds identified. Best-match compounds for each retention time and replicate tube were then chosen based on Net match score. Peak areas of any compounds identified in blank control tubes were removed from the peak areas of all samples within a GC-MS run. Missing values for compounds (i.e. those compounds found only in some technical replicate tubes) were zero-filled.

### 4.3 Statistics and data visualisation for volatile analysis

For all VOC data (Figure 2, Figure 3), TIC (total ion count) data for all compounds was converted to relative amount (%) using the formula ion count/total ion count of sample.

For the single (November 2024) snapshot, multivariate analysis were performed using all 245 retained compounds following post-processing and filtering. Compound abundance data were square-root transformed to reduce the influence of highly abundant compounds and to mitigate zero inflation, following established ecological recommendations (Legendre and Legendre 2012; 2013; Chang and Lin 2011).

For Principal Component Analysis (PCA, Figure 2A) transformed data were mean-centred and scaled to unit variance to ensure comparability among compounds with differing absolute abundances. PCA was conducted using the FactoMineR package with visualisation provided by factoextra in R (Kassambara and Mundt 2017; Lê, Josse, and Husson 2008). To assess compositional differences in VOC profiles among sampling locations, a permutational multivariate analysis of variance (PERMANOVA) was performed using the adonis2 function from the vegan R package, based on Euclidean distances and 9,999 permutations (Oksanen 2015).

Hierarchical Cluster Analysis (HCA, Figure 2B) was conducted using Ward’s minimum variance linkage method with Euclidean distance, a combination that favours the formation of compact and internally coherent clusters. Cluster structure identified by HCA was subsequently used to define groups for downstream exploratory analyses. Partial Least Squares Discriminant Analysis (PLS-DA) was applied as a supervised exploratory method to characterise compounds contributing most strongly to differences among the three major clusters identified by HCA. PLS-DA was implemented using the pls and mixOmics R packages (Rohart et al. 2017). To minimise over-interpretation, PLS-DA results were not used for hypothesis testing but to identify patterns in variable contributions. The top 10 compounds with the highest Variable Importance in Projection (VIP) scores for each cluster are reported in Table 1.

**Table 1:**
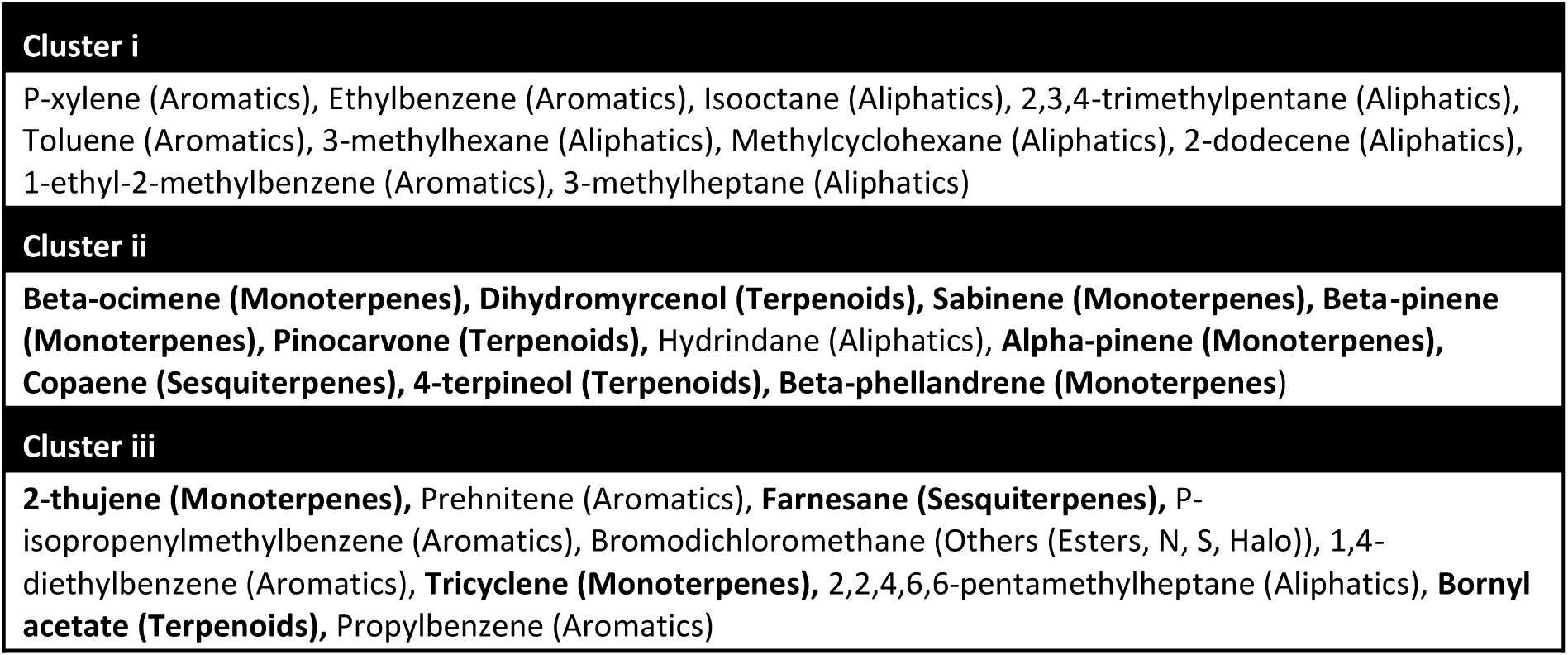
Top 10 compounds from PLS-DA VIP analysis (partial least squares discriminant analysis using variable importance in projection) of the first 3 clusters of the HCA (hierarchical cluster analysis) analysis in Figure 2B: Bold compounds highlight terpenes/terpenoids.

To examine differences in VOC composition using ecologically interpretable groupings, compounds were classified into six functional classes: aliphatics, aromatics, monoterpenes, sesquiterpenes, terpenoids, and others (including esters and nitrogen- and sulphur-containing compounds) (Figure 2C and D). For each class, compound richness (number of compounds) and summed relative abundance were calculated per location. One-way ANOVAs were used to test for differences among locations, with post-hoc Tukey’s HSD tests applied to identify pairwise differences in both summed abundance (Table S 2 & Table S 3) and compound richness (Table S 4 & Table S 5).

Spatial variation in terpene abundance was visualised using a hierarchical heatmap (Figure 2E) in which colour intensity represents compound abundance scaled within individual locations.

To contextualise VOC profiles in relation to anthropogenic influence, the relative abundance of all terpenes and terpenoids was expressed as a ratio to the summed relative abundance of anthropogenic BTEX compounds (Figure 2F, benzene, toluene, ethylbenzene, and p-xylene; ∑terpenes/∑BTEX), following established approaches for assessing biogenic versus anthropogenic dominance (Malik et al. 2025).

For analysis of VOC profiles over time in the single sampling site (Figure 3), relative amount data was used. The average relative amounts of all identified biogenic volatiles were summed within six compound classes and compared across the 7 time-points.

All statistics were run using R version 4.4.1 (2024-06-14 ucrt) within RStudio 2024.09.0+375 “Cranberry Hibiscus” release for windows. All graphs were generated using the ggplot2 (Wickham 2011) package.

### 4.4 Statistical modelling of meteorological drivers of bVOC emissions

To investigate environmental drivers of bVOC emissions within a single greenspace (Botanical Gardens Outdoors) across the calendar year, linear mixed-effects models were fitted to compound class–level emission data from the Oxford Botanic Garden Outdoor site (BO) using R (v.4.x). Peak areas were averaged across technical replicates for each sampling date prior to analysis. Compounds were grouped into five functional classes: monoterpenes, sesquiterpenes, terpenoids, aromatics, aliphatics, and others (esters, nitrogen-, sulphur- and halogen-containing compounds), and summed abundances were calculated for each class per sampling date. Response variables were log-transformed using log(1 + x) to stabilise variance and reduce right skew. Predictor variables included mean daily temperature (°C), relative humidity (%), daily precipitation (mm), and wind speed (m s⁻¹). All predictors were centred and scaled (z-score transformation) prior to modelling to facilitate comparison of effect sizes. For each compound class, a linear mixed-effects model was fitted using the lme4 package, with sampling date included as a random intercept to account for repeated measurements over time:

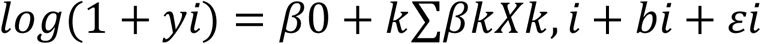

where 𝑦_𝑖_is the summed abundance of a given compound class on sampling date 𝑖, 𝑋_𝑘_are scaled meteorological predictors, 𝑏_𝑖_is the random effect of sampling date, and 𝜀_𝑖_is the residual error.

Statistical significance of fixed effects was assessed using Wald tests implemented via the lmerTest package. Model coefficients and standard errors were extracted for visualisation as coefficient plots and heatmaps, with colour intensity representing the direction and magnitude of standardised effects. Model structure was defined a priori based on hypothesised meteorological drivers, and model fit was assessed to ensure consistency across compound classes.

## 5. Results

### 5.1 A snapshot in time: distinctive fingerprints of six urban greenspaces

Our analysis pipeline resulted in 245 unique compounds being identified within our 6 Oxford sampling sites (Table S 1). A PCA (Figure 2A) using all identified compounds as predictor variables revealed significant site-specific clustering (PERMANOVA, F5,16 = 8.889, p = 0.001) with pair-wise separation of all sites (p < 0.05) except Wytham Woods (WY) in the Warneford Meadow (WA, PERMANOVA, p = 0.846). Further HCA analysis further revealed site-level structuring of most VOC profiles (Figure 2B). Primarily, University Parks Open (UO) and Botanical Gardens Outdoors (BO) separate from all other sites (Figure 2B Cluster i). PLS-DA VIP analysis (Table 1 and Table S7), due to higher amounts of many anthropogenic emission-related compounds such as 2,3,3-trimethylpentane, p-xylene, 2-ethylhexan-1-ol, indane, and ethylbenzene. Plant related compounds (beta-ocimene, dihydromyrcenol, sabinene, diacetone alcohol, beta-pinene, pinocarvone) were chiefly responsible for the secondary clustering (Figure 2B Cluster ii) due to presence (or higher proportions) in the Rainforest Glasshouse (RH) compared with University Parks Dense (UD), Wytham Woods (WY), and Warneford Meadow (WA). The third most significant cluster (Figure 2B Cluster iii) splits University Parks Open (UO) and Botanical Gardens Outdoors (BO), again due to the higher proportion/presence of terpenes/terpenoids such as 2-thujene, farnesane, and bornyl acetate found in Botanical Gardens Outdoors. Full results of PLS-DA analysis can be found in Table S7.

#### 5.1.1 Urban greenspaces show distinctive mix of compound classes

The number of identified terpenoids, monoterpenes and sesquiterpenes was found to be highest in the Rainforest Glasshouse (13, 15, 10 respectively). These compound numbers were significantly higher than the other 5 sites (one-way ANOVAs followed by Tukey’s HSD post-hoc test p < 0.05, Figure 2D - full statistics in Table S 4 & Table S 5).Numerically, the Botanical Gardens Outdoors was the outdoor location with the highest counts of terpenoids, monoterpenes and sesquiterpenes (8, 13, 2 respectively), closely followed by Wytham (9, 12, 1) In terms of relative abundance (Figure 2C - full statistics in Table S 2 & Table S 3), Botanic Garden Outdoors contained a significantly higher proportion of aromatic compounds compared to other sites suggesting a strong influence from anthropogenic sources. The Rainforest Glasshouse showed significantly higher proportion of monoterpenes, whereas sesquiterpenes were shared by the Rainforest Glasshouse and Botanical Gardens Outdoors (One-way ANOVAs followed by Tukey’s HSD post-hoc test p < 0.05).

#### 5.1.2 Presence/absence of plant-based terpenes and terpenoids

A total of 54 terpenes/terpenoids were identified in this work (Figure 2G). Ten compounds - 1-8 cineole (eucalyptol), 3-carene, alpha-calacorene, alpha-pinene, beta-pinene, camphene, camphor, limonene, menthol, and p-cymene - were detected at every site, with a further two – menthone and tricyclene – detected in five of the six sampling sites. Again, the Rainforest Glasshouse exhibited the largest number of identified terpenes/terpenoid compounds overall (38 compounds) of which 16 were not detected at other sites. Of the outdoor sites, Botanical Gardens Outdoors contained the highest number of identified terpenes/terpenoid (23), of which 6 were uniquely present at this site. University Parks Dense (17), University Parks Open (14), Warneford Meadow (14) and Wytham Forest (22) all showed an overlapping but distinctive terpene mix. A full breakdown of compounds unique to individual sites is shown in Table S 6.

#### 5.1.3 BTEX compounds as predictors of pollution

The ratio of biogenic terpenes/terpenoids to anthropogenic BTEX compounds was significantly higher in the Rainforest Glasshouse compared to the other sites (Figure 2E, one-way ANOVA followed by Tukey’s HSD post-hoc test p < 0.001). This is in part due to the increased prevalence of terpenes/terpenoids and in part to the lower proportion of BTEX compounds (Figure 2F) in this indoor space. The outdoor sites all had a significantly lower ratios but were not significantly different from each other.

### 5.2 How do urban greenspace bVOC profiles vary through time under real-world conditions?

Using the same classification system for our identified compounds we counted both compound numbers and summed abundances to visualise each class over time. In terms of the relative abundances, all compound classes peaked during late spring to summer (May to August 2025) with two notable outliers - monoterpenes had an early peak on November 27^th^, 2024, and terpenoids had a late peak on October 29^th^, 2025. Interestingly, in terms of the absolute number of compounds identified, it was early spring (04 March 2025) which showed peaks for aliphatics (42), aromatics (14), terpenoids (10) and others (9), with monoterpene counts being more stable over the year.

Mixed-effects modelling revealed that different components of the biogenic scentscape responded in contrasting ways to short-term environmental variation (Figure 3C). Temperature emerged as the most consistent positive driver of terpene emissions, with higher temperatures associated with increased relative abundances of both monoterpenes and sesquiterpenes. Humidity also showed compound-class-specific effects with terpenes exhibiting a significant positive association with increased levels of relative humidity, whereas aromatic and aliphatic compounds were weaker or neutral. Increased wind speed contributed to increases in both aromatic and aliphatic fractions.

## 6. Discussion

### 6.1 bVOCs and aVOCs shape urban greenspace scentscapes

An initial PCA of our six sampling sites on a single date (Nov 27^th^, 2024) revealed significant site-specific clustering. Further HCA revealed site structuring was due to both bVOCs and aVOCs (Table 1, Figure 2B). We chose to interrogate the first 3 clusters which appeared. The first cluster separated two outdoor sites - Botanical Gardens Outdoors and University Parks Open – from all other sampling sites. PLS-DA VIP analysis showed that this was due to many emission-related aVOCs (Table 1) including the traffic-related BTEX compounds (Figure 2F). These two sites are very close to a busy city centre roads (Figure 1A) suggesting that traffic is the likely main contributor. This is supported by BTEX presence measurements where over 40% of identified volatiles were BTEX compounds in the Botanical Gardens Outdoors and University Parks Open, but just 13% in Wytham Woods and Warneford Meadow which are more isolated from traffic. The relationship between proximity to traffic and BTEX prevalence has not been assessed in this paper but has been previously reported (Hoque et al. 2008).

Interestingly, despite its proximity to University Parks Open (∼80 m), University Parks Dense showed markedly lower BTEX levels (43% to 29%). Localised differences in BTEX presence within urban greenspaces has been demonstrated before; for example, Upmanis et al. (2001) reported that concentrations of benzene and toluene were up to one-third lower just 40m into urban parkland within two large Scandinavian cities. A second study also showed decreases in particulate matter (PM10 and PM2.5) of up to 50% at 200m (Gómez-Moreno et al. 2019). Additionally, the presence of exposed soil and leaf litter layer around trees in University Parks Dense compared to the grassy University Parks Open site may contribute to increased degradation of BTEX compounds through microbial metabolism. A 2012 study, for example, showed that by adding a mixed bacterial consortium to soil, BTEX compounds were more rapidly degraded (Mukherjee and Bordoloi 2012).

Biogenic volatiles, particularly terpenes (Table 1, Figure 2G), were chiefly responsible for the secondary clustering of samples from Rainforest House, and those of Wytham Woods and Warneford Meadow. In total across all sampling sites, we identified 54 unique terpenes/terpenoids. The Rainforest Glasshouse contained the highest number of terpenes (38) with considerable spread across the outdoor sampling sites (14 to 23). The presence of these compounds is highly suggestive of multiple and specific health benefits within each location. For example, 10 terpenes/terpenoids were ubiquitous across all sampling sites - compounds which have also been found in air samples elsewhere using similar methods (Gu et al. 2024; Bryant et al. 2022; Walker et al. 2023; Hellén et al. 2024; Jones, Hopkins, and Lewis 2011; Hakola et al. 2003; Detournay et al. 2011). These compounds alone are cited to have anti-inflammatory, anxiolytic, antimicrobial, immune-boosting, antidepressant, anaesthetic, gastroprotective, antioxidant, anti-cancer (*in vitro*), immune-modulatory, sedative, hypolipidemic, analgesic, decongestant, bronchodilatory, mucolytic, and cognitive enhancing effects (Lee et al. 2022; Vallianou and Hadzopoulou-Cladaras 2016; Salehi et al. 2019; Oliveira et al. 2024; Sun 2007; Woo et al. 2019; Jo et al. 2021). It could be argued, therefore, that spending time in any of these sites could impart significant health gains. However, the relationships between these compounds and their benefits are likely concentration dependent, therefore further methodological work is needed to quantify and make precise recommendations such as when, and for how long, to visit specific greenspaces. Previously it was found that just 20 minutes is adequate to derive physiological and psychological benefits from time in woodland (Haluza et al. 2025; Park et al. 2010).

Another useful metric to separate our sites is a ratio of biogenic to anthropogenic compounds. An example of this can be found in study which assessed air over Covid lockdown in 2020 at an urban site in India (Malik et al. 2025) in which a sharp increase in the ∑α+beta--pinene/∑BTEX ratios during the strict lockdown phase was hypothesised to be due to a reduction in traffic related activity. We mirrored this study and found that, on a particular day, the Rainforest House had significantly higher terpene/BTEX ratio than all the other sites (Figure 2F). This result is likely due to the interplay of two different factors known to affect the volatile makeup of indoor spaces. On the one hand, this glasshouse is plant rich and enclosed and as such likely acts as an enclosure for those bVOCs produced within, and conversely, aVOC ingression may be limited. These results suggest that plant rich greenhouses may be the most beneficial place to spend time, however, in a 2006-2007 study, aVOCs were found to be significantly higher in indoor sites than outdoor sites in the vicinity of an industrial park (Chang et al. 2019), showing that simply being inside is little protection from external sources of pollution.

### 6.2 The impacts of seasonality on bVOC emissions in the Botanical Gardens

As well as looking at a snapshot of the scentscapes across the six urban greenspaces, we also explored how bVOC change across time and in the face of environmental variation in the Botanical Outdoors location. Using the same classification system for our identified compounds we counted both compound numbers and summed abundances to visualise each class over time. In terms of the relative abundances, all compound classes peaked during late spring to summer (May to August 2025) with two notable outliers - monoterpenes had an early peak on November 27^th^, 2024, and terpenoids had a late peak on October 29^th^, 2025. Interestingly, in terms of the absolute number of compounds identified, it was early spring (March 4th) which showed peaks for aliphatics (42), aromatics (14), terpenoids (10) and others (9), with monoterpene counts being more stable over the year.

Higher temperature and relative humidity were the most consistent positive predictors of biogenic emissions, particularly for terpenes. For temperature this is likely linked to the well-established positive relationship between emission rates and leaf temperatures via both biosynthesis through increased enzymatic activity and volatilisation vapour pressure (Guenther et al. 1993; Luo et al. 2025). Aliphatics also showed a positive temperature response, likely driven by enhanced volatilisation at higher temperatures but also resistance to biodegradation (Abbasian et al. 2015). In contrast, terpenoids, which are oxygenated terpenes and much more reactive, declined with increasing temperature. This is consistent with their greater susceptibility to photochemical degradation at higher temperatures (Atkinson and Arey 2003; Hallquist et al. 2009). The role of humidity in terpene/terpenoid presence may be linked to an increase in stomatal opening and subsequent environmental release when relative humidity is higher (Arve and Torre 2015).

Precipitation effects also differed among VOC classes. Terpene abundances showed a positive association with precipitation. Wetting is known to give a short-term pulse release through mechanical displacement from soil air spaces after rain (Pugliese et al. 2023) and this was likely the reason for the high amount recorded on November 27^th^, 2024, where 8.10 mm of rain was recorded. In contrast, aromatics and aliphatics showed a negative association with precipitation on the day of sampling, consistent with atmospheric dilution, wet deposition, or reduced resuspension of background hydrocarbons rather than changes in emission strength (Casas et al. 2021). There was a second rainfall event on September 4^th^, 2025 (9.4 mm), however, this followed 16.3 mm of rain on the previous day which is likely to have negatively affected soil-based terpene release on the day of sampling. Notably, only two sampling events coincided with measurable rainfall so these patterns need further exploration in urban greenspaces.

Wind speed effects showed significant positive associations for aromatic and aliphatic compounds. Given their relatively long atmospheric lifetimes compared to terpenes and terpenoids (Luo et al. 2025; Hellén et al. 2018), these increases are more likely to reflect regional background concentrations than increases in local emissions. In contrast, terpene emissions showed little sensitivity to wind speed, consistent with their predominantly local, vegetation-derived sources and rapid atmospheric reactivity leading to degradation if produced distantly.

Overall, these results indicate that temperature, precipitation, wind speed and humidity all impact on the detection of different bVOC chemical classes. The absence of a single dominant predictor across all VOC classes highlights the importance of considering scentscapes as multidimensional chemical systems, shaped by interacting biological and meteorological processes. However, this class-specific sensitivity to environmental drivers adds understanding of when and where urban greenspaces may be most chemically beneficial for human exposure. These results are suggestive that warm, humid, and (unfortunately) rainy conditions increase the levels of health-promoting terpene/terpenoid compounds in the air and as such are likely the most beneficial conditions for humans to spend time in urban greenspaces.

### 6.3 Methodological advancements, limitations, and future work

As well as providing an understanding of urban greenspace scentscapes over time, this study aimed to develop a robust and repeatable protocol for measuring the presence and relative amounts of a wide range of volatiles of interest. Although similar methods have been developed for headspace sampling (Yin et al. 2022; de Carvalho Couto et al. 2024; Reyrolle et al. 2024), air sampling studies have historically employed targeted approaches that significantly limit the number of compounds identified. For instance, previous studies have used between 15 and 66 VOCs across various forest and urban environments (Walker et al. 2023; Pripdeevech et al. 2025; Mula et al. 2024). In contrast, our study adopted an untargeted metabolomics-style approach commonly used in biological matrices to enhance the depth and resolution of VOC profiling (Fernie et al. 2011; Van den Berg et al. 2006; Rosenthal et al. 2024) to identify 245 unique compounds, of which 54 were unique terpenes/terpenoids. This is significantly higher than any studies on ambient air so far published (Walker et al. 2023; Sanaei et al. 2023; Mula et al. 2024). This improved identification capacity is likely attributable to several key enhancements in our analytical pipeline: greater technical replication, the use of a Kováts retention index, a more focused compound library, a slow GC temperature ramp, and the use of deconvolution for resolving co-eluting peaks. Furthermore, the tight clustering of technical replicates observed in the PCA (Figure 2A) confirms that our collection methods and downstream processing are sufficiently controlled to ensure analytical reproducibility and minimise technical variability.

Despite these improvements, our analyses capture only a fraction of the chemical complexity present. In most samples we detect far more chromatographic features than we can confidently identify, and the ∼245 compounds reported here represent ∼10% (or less) of the total compounds present. This limitation is expected in untargeted ambient-air GC–MS, where a large proportion of peaks correspond to (i) compounds absent from reference libraries, (ii) compounds lacking retention index (RI) values on comparable stationary phases, (iii) low-abundance or highly reactive species near the detection limit, and (iv) co-eluting mixtures that remain partially unresolved even after deconvolution. As a result, our interpretation focuses on the “identifiable fraction” of the scentscape rather than the full atmospheric mixture. One major benefit of TD work is the ability for the ‘recollection’ of the split portion of the GC-MS analysis. Future work intends to use this feature to analyse the same samples on multiple GC column types (polar/non-polar) – this will help to resolve significantly more atmospheric compounds.

Identification confidence is a further constraint. We used a minimum Net match threshold of 70, which is permissive and may increase the risk of imperfect annotations, particularly for isomeric compounds and low-intensity peaks. Although 99 compounds had a match factor > 90, the mean match factor for retained identifications was 86. Therefore, a subset of compounds should be regarded as putative with future work concentrating on targeted confirmation of key marker compounds using standards. Our identifications are strengthened by combining spectral matching with RI matching, however, expanding the RI coverage of our library on DB-5–type phases and continuing to develop an in-house “Oxford greenspace” standard set would improve annotation rates. Additionally, we did not employ chiral separation in this work, meaning stereoisomers of compounds such as Alpha-pinene (+/-) were not resolved. These may be important for understanding biological source and ecological function and as such could be included in future. Finally, to truly overcome co-elution, two-dimensional gas chromatography (GC x GC) could be used to separate overlapping peaks without the need for software-based deconvolution.

Our sampling design also imposes important constraints. First, sampling was conducted within a narrow daytime window (11:00–13:00). As our models show, many bVOCs are affected by environmental changes, therefore, abundance and composition likely differ through the day. Second, we used a fixed sampling duration (2 h) and flow rate (100 ml/min). This equates to just 2 minutes of human breathing. Longer sampling times or higher sampled volumes would likely increase detection of low-abundance compounds, but at the risk of sorbent saturation, breakthrough of other volatile species, and increased influence meteorology during the sampling window. Third, while technical replication was employed, biological/environmental replication was limited: the “snapshot” comparison is based on a single calendar date, and the year-long analysis focuses on one site. In both cases, sampling was carried out at a precise location within each site. As such, our site-level inferences should not be interpreted as definitive rankings of whole sites across seasons and years.

Quantification is another limitation. Relative measures are useful for compositional comparisons, but absolute quantification will be essential to translate scentscape metrics into health-relevant exposure estimates. The quantification of compounds such as terpenes will help to support policy-facing recommendations.

Finally, the environmental predictors included here (temperature, humidity, precipitation, wind speed) are only a subset of the drivers that shape ambient VOC fingerprints. A more holistic predictive framework will need to incorporate additional covariates such as (i) local vegetation composition and phenology (biodiversity and functional traits), (ii) microclimate at the sampling point (solar radiation, canopy cover, boundary-layer stability, VPD), (iii) soil moisture and microbial activity, (iv) episodic disturbances (mowing, leaf litter pulses, pest/pathogen outbreaks), and (v) urban pollution context (traffic intensity, background NOx/O₃, and atmospheric oxidation capacity). Incorporating these variables—alongside expanded temporal coverage, multi-year replication, and partial targeted quantification—will be necessary to move from descriptive classification of scentscapes to robust prediction of when and where greenspaces maximise health-relevant exposures.

## Data Availability

Raw AGILENT files are available here: http://doi.org/10.5281/zenodo.18493789

## 7. Author Contributions

William Kay was responsible for methods development, data collection and all data analyses. William Kay and Anya Lindstrom Battle were both involved in the writing and editing of the manuscript. Kathy Willis conceived of the initial ideas and methodologies and was also involved in the writing and editing of the manuscript. Mahal Humberstone, Molly Tucker and Kieran Storer all contributed to in-field sample collection. Geoffrey Kite was involved in the GC-MS data collection, as well as providing useful guidance. All authors contributed critically to the drafts and gave final approval for publication.

## 9. Data Availability Statement

Raw AGILENT files are available here: http://doi.org/10.5281/zenodo.18493789

## 10. Acknowledgments

We would firstly like to thank the University of Oxford Botanic Garden and Wytham Woods for permission to sample on their sites. We would also like to thank Dr Ben Martin who provided helpful advice for sampling and analysis as well as the development of early sampling methodologies, and Dr Luciana Carvalho who always had time to share thoughts and ideas relating to the project. In addition, we thank Dr Pedro Bota for his generosity of time, availability and equipment. Finally, we thank all members of the Oxford long-term ecology lab for their ongoing feedback.

## 13. Supplementary Information

**Table S 1:**
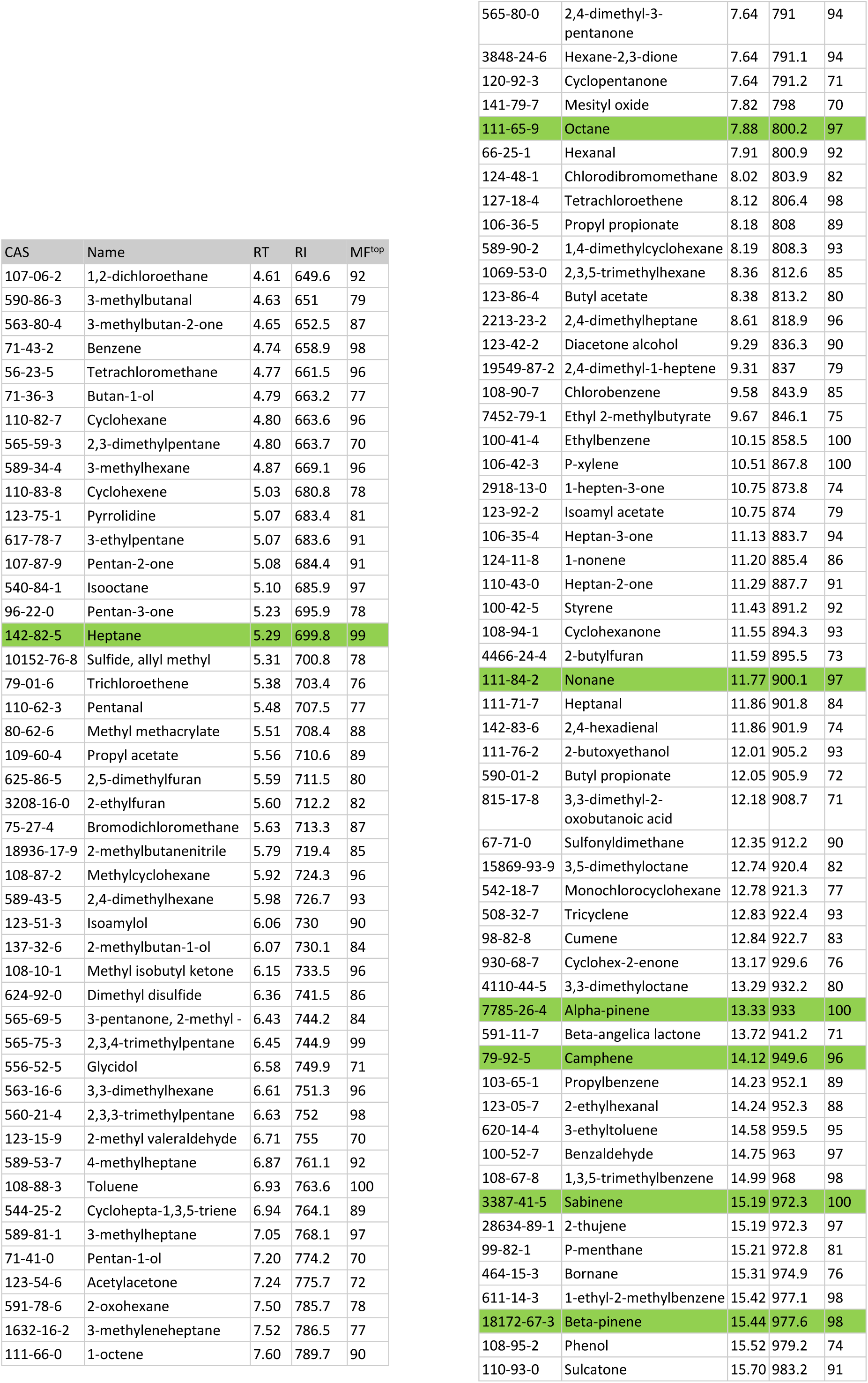
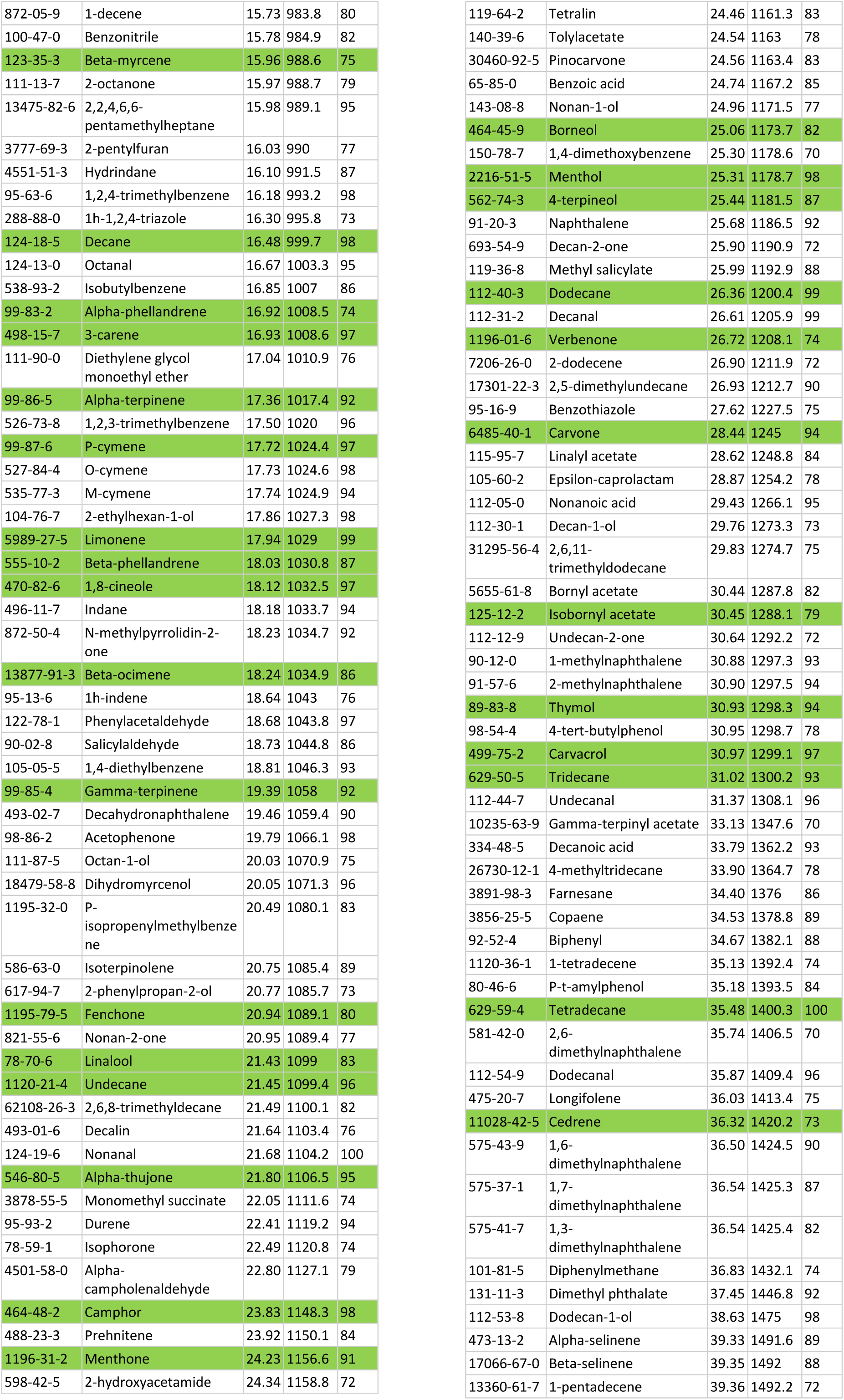
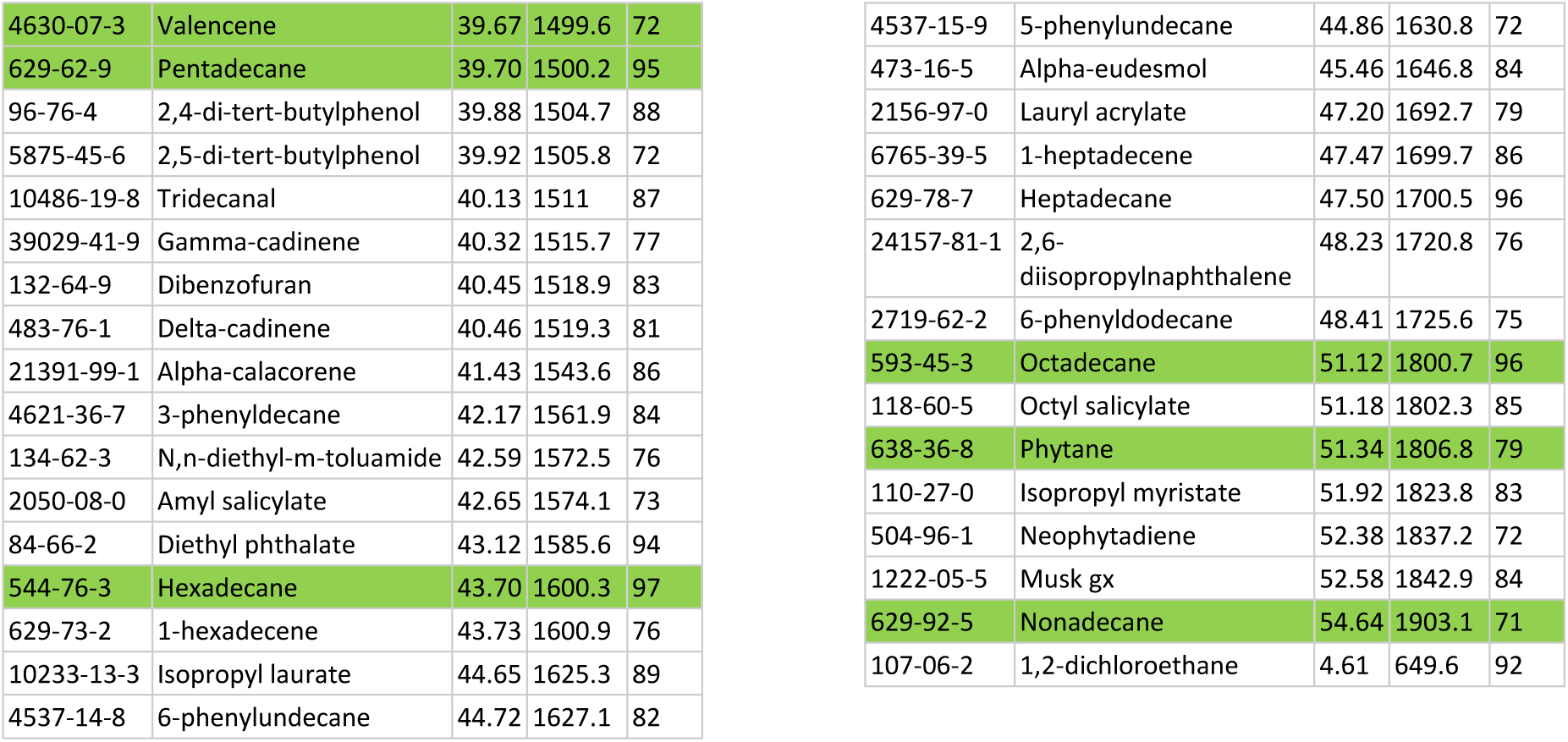
A full list of the compounds detected on 27^th^ November 2024. List is sorted by retention time (RT). RI is retention index values corresponding to Kovats index. MF^top^ is the highest match factor found over all samples. Green rows show compounds for which we have confirmed RT via standard solutions.

| CAS | Name | RT | RI | MF<sup>top</sup> |
| --- | --- | --- | --- | --- |
| 107-06-2 | 1,2-dichloroethane | 4.61 | 649.6 | 92 |
| 590-86-3 | 3-methylbutanal | 4.63 | 651 | 79 |
| 563-80-4 | 3-methylbutan-2-one | 4.65 | 652.5 | 87 |
| 71-43-2 | Benzene | 4.74 | 658.9 | 98 |
| 56-23-5 | Tetrachloromethane | 4.77 | 661.5 | 96 |
| 71-36-3 | Butan-1-ol | 4.79 | 663.2 | 77 |
| 110-82-7 | Cyclohexane | 4.80 | 663.6 | 96 |
| 565-59-3 | 2,3-dimethylpentane | 4.80 | 663.7 | 70 |
| 589-34-4 | 3-methylhexane | 4.87 | 669.1 | 96 |
| 110-83-8 | Cyclohexene | 5.03 | 680.8 | 78 |
| 123-75-1 | Pyrrolidine | 5.07 | 683.4 | 81 |
| 617-78-7 | 3-ethylpentane | 5.07 | 683.6 | 91 |
| 107-87-9 | Pentan-2-one | 5.08 | 684.4 | 91 |
| 540-84-1 | Isooctane | 5.10 | 685.9 | 97 |
| 96-22-0 | Pentan-3-one | 5.23 | 695.9 | 78 |
| 142-82-5 | Heptane | 5.29 | 699.8 | 99 |
| 10152-76-8 | Sulfide, allyl methyl | 5.31 | 700.8 | 78 |
| 79-01-6 | Trichloroethene | 5.38 | 703.4 | 76 |
| 110-62-3 | Pentanal | 5.48 | 707.5 | 77 |
| 80-62-6 | Methyl methacrylate | 5.51 | 708.4 | 88 |
| 109-60-4 | Propyl acetate | 5.56 | 710.6 | 89 |
| 625-86-5 | 2,5-dimethylfuran | 5.59 | 711.5 | 80 |
| 3208-16-0 | 2-ethylfuran | 5.60 | 712.2 | 82 |
| 75-27-4 | Bromodichloromethane | 5.63 | 713.3 | 87 |
| 18936-17-9 | 2-methylbutanenitrile | 5.79 | 719.4 | 85 |
| 108-87-2 | Methylcyclohexane | 5.92 | 724.3 | 96 |
| 589-43-5 | 2,4-dimethylhexane | 5.98 | 726.7 | 93 |
| 123-51-3 | Isoamylol | 6.06 | 730 | 90 |
| 137-32-6 | 2-methylbutan-1-ol | 6.07 | 730.1 | 84 |
| 108-10-1 | Methyl isobutyl ketone | 6.15 | 733.5 | 96 |
| 624-92-0 | Dimethyl disulfide | 6.36 | 741.5 | 86 |
| 565-69-5 | 3-pentanone, 2-methyl - | 6.43 | 744.2 | 84 |
| 565-75-3 | 2,3,4-trimethylpentane | 6.45 | 744.9 | 99 |
| 556-52-5 | Glycidol | 6.58 | 749.9 | 71 |
| 563-16-6 | 3,3-dimethylhexane | 6.61 | 751.3 | 96 |
| 560-21-4 | 2,3,3-trimethylpentane | 6.63 | 752 | 98 |
| 123-15-9 | 2-methyl valeraldehyde | 6.71 | 755 | 70 |
| 589-53-7 | 4-methylheptane | 6.87 | 761.1 | 92 |
| 108-88-3 | Toluene | 6.93 | 763.6 | 100 |
| 544-25-2 | Cyclohepta-1,3,5-triene | 6.94 | 764.1 | 89 |
| 589-81-1 | 3-methylheptane | 7.05 | 768.1 | 97 |
| 71-41-0 | Pentan-1-ol | 7.20 | 774.2 | 70 |
| 123-54-6 | Acetylacetone | 7.24 | 775.7 | 72 |
| 591-78-6 | 2-oxohexane | 7.50 | 785.7 | 78 |
| 1632-16-2 | 3-methyleneheptane | 7.52 | 786.5 | 77 |
| 111-66-0 | 1-octene | 7.60 | 789.7 | 90 |
| 565-80-0 | 2,4-dimethyl-3-pentanone | 7.64 | 791 | 94 |
| 3848-24-6 | Hexane-2,3-dione | 7.64 | 791.1 | 94 |
| 120-92-3 | Cyclopentanone | 7.64 | 791.2 | 71 |
| 141-79-7 | Mesityl oxide | 7.82 | 798 | 70 |
| 111-65-9 | Octane | 7.88 | 800.2 | 97 |
| 66-25-1 | Hexanal | 7.91 | 800.9 | 92 |
| 124-48-1 | Chlorodibromomethane | 8.02 | 803.9 | 82 |
| 127-18-4 | Tetrachloroethene | 8.12 | 806.4 | 98 |
| 106-36-5 | Propyl propionate | 8.18 | 808 | 89 |
| 589-90-2 | 1,4-dimethylcyclohexane | 8.19 | 808.3 | 93 |
| 1069-53-0 | 2,3,5-trimethylhexane | 8.36 | 812.6 | 85 |
| 123-86-4 | Butyl acetate | 8.38 | 813.2 | 80 |
| 2213-23-2 | 2,4-dimethylheptane | 8.61 | 818.9 | 96 |
| 123-42-2 | Diacetone alcohol | 9.29 | 836.3 | 90 |
| 19549-87-2 | 2,4-dimethyl-1-heptene | 9.31 | 837 | 79 |
| 108-90-7 | Chlorobenzene | 9.58 | 843.9 | 85 |
| 7452-79-1 | Ethyl 2-methylbutyrate | 9.67 | 846.1 | 75 |
| 100-41-4 | Ethylbenzene | 10.15 | 858.5 | 100 |
| 106-42-3 | P-xylene | 10.51 | 867.8 | 100 |
| 2918-13-0 | 1-hepten-3-one | 10.75 | 873.8 | 74 |
| 123-92-2 | Isoamyl acetate | 10.75 | 874 | 79 |
| 106-35-4 | Heptan-3-one | 11.13 | 883.7 | 94 |
| 124-11-8 | 1-nonene | 11.20 | 885.4 | 86 |
| 110-43-0 | Heptan-2-one | 11.29 | 887.7 | 91 |
| 100-42-5 | Styrene | 11.43 | 891.2 | 92 |
| 108-94-1 | Cyclohexanone | 11.55 | 894.3 | 93 |
| 4466-24-4 | 2-butylfuran | 11.59 | 895.5 | 73 |
| 111-84-2 | Nonane | 11.77 | 900.1 | 97 |
| 111-71-7 | Heptanal | 11.86 | 901.8 | 84 |
| 142-83-6 | 2,4-hexadienal | 11.86 | 901.9 | 74 |
| 111-76-2 | 2-butoxyethanol | 12.01 | 905.2 | 93 |
| 590-01-2 | Butyl propionate | 12.05 | 905.9 | 72 |
| 815-17-8 | 3,3-dimethyl-2-oxobutanoic acid | 12.18 | 908.7 | 71 |
| 67-71-0 | Sulfonyldimethane | 12.35 | 912.2 | 90 |
| 15869-93-9 | 3,5-dimethyloctane | 12.74 | 920.4 | 82 |
| 542-18-7 | Monochlorocyclohexane | 12.78 | 921.3 | 77 |
| 508-32-7 | Tricyclene | 12.83 | 922.4 | 93 |
| 98-82-8 | Cumene | 12.84 | 922.7 | 83 |
| 930-68-7 | Cyclohex-2-enone | 13.17 | 929.6 | 76 |
| 4110-44-5 | 3,3-dimethyloctane | 13.29 | 932.2 | 80 |
| 7785-26-4 | Alpha-pinene | 13.33 | 933 | 100 |
| 591-11-7 | Beta-angelica lactone | 13.72 | 941.2 | 71 |
| 79-92-5 | Camphene | 14.12 | 949.6 | 96 |
| 103-65-1 | Propylbenzene | 14.23 | 952.1 | 89 |
| 123-05-7 | 2-ethylhexanal | 14.24 | 952.3 | 88 |
| 620-14-4 | 3-ethyltoluene | 14.58 | 959.5 | 95 |
| 100-52-7 | Benzaldehyde | 14.75 | 963 | 97 |
| 108-67-8 | 1,3,5-trimethylbenzene | 14.99 | 968 | 98 |
| 3387-41-5 | Sabinene | 15.19 | 972.3 | 100 |
| 28634-89-1 | 2-thujene | 15.19 | 972.3 | 97 |
| 99-82-1 | P-menthane | 15.21 | 972.8 | 81 |
| 464-15-3 | Bornane | 15.31 | 974.9 | 76 |
| 611-14-3 | 1-ethyl-2-methylbenzene | 15.42 | 977.1 | 98 |
| 18172-67-3 | Beta-pinene | 15.44 | 977.6 | 98 |
| 108-95-2 | Phenol | 15.52 | 979.2 | 74 |
| 110-93-0 | Sulcatone | 15.70 | 983.2 | 91 |
| 872-05-9 | 1-decene | 15.73 | 983.8 | 80 |
| 100-47-0 | Benzonitrile | 15.78 | 984.9 | 82 |
| 123-35-3 | Beta-myrcene | 15.96 | 988.6 | 75 |
| 111-13-7 | 2-octanone | 15.97 | 988.7 | 79 |
| 13475-82-6 | 2,2,4,6,6-pentamethylheptane | 15.98 | 989.1 | 95 |
| 3777-69-3 | 2-pentylfuran | 16.03 | 990 | 77 |
| 4551-51-3 | Hydrindane | 16.10 | 991.5 | 87 |
| 95-63-6 | 1,2,4-trimethylbenzene | 16.18 | 993.2 | 98 |
| 288-88-0 | 1h-1,2,4-triazole | 16.30 | 995.8 | 73 |
| 124-18-5 | Decane | 16.48 | 999.7 | 98 |
| 124-13-0 | Octanal | 16.67 | 1003.3 | 95 |
| 538-93-2 | Isobutylbenzene | 16.85 | 1007 | 86 |
| 99-83-2 | Alpha-phellandrene | 16.92 | 1008.5 | 74 |
| 498-15-7 | 3-carene | 16.93 | 1008.6 | 97 |
| 111-90-0 | Diethylene glycol monoethyl ether | 17.04 | 1010.9 | 76 |
| 99-86-5 | Alpha-terpinene | 17.36 | 1017.4 | 92 |
| 526-73-8 | 1,2,3-trimethylbenzene | 17.50 | 1020 | 96 |
| 99-87-6 | P-cymene | 17.72 | 1024.4 | 97 |
| 527-84-4 | O-cymene | 17.73 | 1024.6 | 98 |
| 535-77-3 | M-cymene | 17.74 | 1024.9 | 94 |
| 104-76-7 | 2-ethylhexan-1-ol | 17.86 | 1027.3 | 98 |
| 5989-27-5 | Limonene | 17.94 | 1029 | 99 |
| 555-10-2 | Beta-phellandrene | 18.03 | 1030.8 | 87 |
| 470-82-6 | 1,8-cineole | 18.12 | 1032.5 | 97 |
| 496-11-7 | Indane | 18.18 | 1033.7 | 94 |
| 872-50-4 | N-methylpyrrolidin-2-one | 18.23 | 1034.7 | 92 |
| 13877-91-3 | Beta-ocimene | 18.24 | 1034.9 | 86 |
| 95-13-6 | 1h-indene | 18.64 | 1043 | 76 |
| 122-78-1 | Phenylacetaldehyde | 18.68 | 1043.8 | 97 |
| 90-02-8 | Salicylaldehyde | 18.73 | 1044.8 | 86 |
| 105-05-5 | 1,4-diethylbenzene | 18.81 | 1046.3 | 93 |
| 99-85-4 | Gamma-terpinene | 19.39 | 1058 | 92 |
| 493-02-7 | Decahydronaphthalene | 19.46 | 1059.4 | 90 |
| 98-86-2 | Acetophenone | 19.79 | 1066.1 | 98 |
| 111-87-5 | Octan-1-ol | 20.03 | 1070.9 | 75 |
| 18479-58-8 | Dihydromyrcenol | 20.05 | 1071.3 | 96 |
| 1195-32-0 | P-isopropenylmethylbenzene | 20.49 | 1080.1 | 83 |
| 586-63-0 | Isoterpinolene | 20.75 | 1085.4 | 89 |
| 617-94-7 | 2-phenylpropan-2-ol | 20.77 | 1085.7 | 73 |
| 1195-79-5 | Fenchone | 20.94 | 1089.1 | 80 |
| 821-55-6 | Nonan-2-one | 20.95 | 1089.4 | 77 |
| 78-70-6 | Linalool | 21.43 | 1099 | 83 |
| 1120-21-4 | Undecane | 21.45 | 1099.4 | 96 |
| 62108-26-3 | 2,6,8-trimethyldecane | 21.49 | 1100.1 | 82 |
| 493-01-6 | Decalin | 21.64 | 1103.4 | 76 |
| 124-19-6 | Nonanal | 21.68 | 1104.2 | 100 |
| 546-80-5 | Alpha-thujone | 21.80 | 1106.5 | 95 |
| 3878-55-5 | Monomethyl succinate | 22.05 | 1111.6 | 74 |
| 95-93-2 | Durene | 22.41 | 1119.2 | 94 |
| 78-59-1 | Isophorone | 22.49 | 1120.8 | 74 |
| 4501-58-0 | Alpha-campholenaldehyde | 22.80 | 1127.1 | 79 |
| 464-48-2 | Camphor | 23.83 | 1148.3 | 98 |
| 488-23-3 | Prehnitene | 23.92 | 1150.1 | 84 |
| 1196-31-2 | Menthone | 24.23 | 1156.6 | 91 |
| 598-42-5 | 2-hydroxyacetamide | 24.34 | 1158.8 | 72 |
| 119-64-2 | Tetralin | 24.46 | 1161.3 | 83 |
| 140-39-6 | Tolylacetate | 24.54 | 1163 | 78 |
| 30460-92-5 | Pinocarvone | 24.56 | 1163.4 | 83 |
| 65-85-0 | Benzoic acid | 24.74 | 1167.2 | 85 |
| 143-08-8 | Nonan-1-ol | 24.96 | 1171.5 | 77 |
| 464-45-9 | Borneol | 25.06 | 1173.7 | 82 |
| 150-78-7 | 1,4-dimethoxybenzene | 25.30 | 1178.6 | 70 |
| 2216-51-5 | Menthol | 25.31 | 1178.7 | 98 |
| 562-74-3 | 4-terpineol | 25.44 | 1181.5 | 87 |
| 91-20-3 | Naphthalene | 25.68 | 1186.5 | 92 |
| 693-54-9 | Decan-2-one | 25.90 | 1190.9 | 72 |
| 119-36-8 | Methyl salicylate | 25.99 | 1192.9 | 88 |
| 112-40-3 | Dodecane | 26.36 | 1200.4 | 99 |
| 112-31-2 | Decanal | 26.61 | 1205.9 | 99 |
| 1196-01-6 | Verbenone | 26.72 | 1208.1 | 74 |
| 7206-26-0 | 2-dodecene | 26.90 | 1211.9 | 72 |
| 17301-22-3 | 2,5-dimethylundecane | 26.93 | 1212.7 | 90 |
| 95-16-9 | Benzothiazole | 27.62 | 1227.5 | 75 |
| 6485-40-1 | Carvone | 28.44 | 1245 | 94 |
| 115-95-7 | Linalyl acetate | 28.62 | 1248.8 | 84 |
| 105-60-2 | Epsilon-caprolactam | 28.87 | 1254.2 | 78 |
| 112-05-0 | Nonanoic acid | 29.43 | 1266.1 | 95 |
| 112-30-1 | Decan-1-ol | 29.76 | 1273.3 | 73 |
| 31295-56-4 | 2,6,11-trimethyldodecane | 29.83 | 1274.7 | 75 |
| 5655-61-8 | Bornyl acetate | 30.44 | 1287.8 | 82 |
| 125-12-2 | Isobornyl acetate | 30.45 | 1288.1 | 79 |
| 112-12-9 | Undecan-2-one | 30.64 | 1292.2 | 72 |
| 90-12-0 | 1-methylnaphthalene | 30.88 | 1297.3 | 93 |
| 91-57-6 | 2-methylnaphthalene | 30.90 | 1297.5 | 94 |
| 89-83-8 | Thymol | 30.93 | 1298.3 | 94 |
| 98-54-4 | 4-tert-butylphenol | 30.95 | 1298.7 | 78 |
| 499-75-2 | Carvacrol | 30.97 | 1299.1 | 97 |
| 629-50-5 | Tridecane | 31.02 | 1300.2 | 93 |
| 112-44-7 | Undecanal | 31.37 | 1308.1 | 96 |
| 10235-63-9 | Gamma-terpinyl acetate | 33.13 | 1347.6 | 70 |
| 334-48-5 | Decanoic acid | 33.79 | 1362.2 | 93 |
| 26730-12-1 | 4-methyltridecane | 33.90 | 1364.7 | 78 |
| 3891-98-3 | Farnesane | 34.40 | 1376 | 86 |
| 3856-25-5 | Copaene | 34.53 | 1378.8 | 89 |
| 92-52-4 | Biphenyl | 34.67 | 1382.1 | 88 |
| 1120-36-1 | 1-tetradecene | 35.13 | 1392.4 | 74 |
| 80-46-6 | P-t-amylphenol | 35.18 | 1393.5 | 84 |
| 629-59-4 | Tetradecane | 35.48 | 1400.3 | 100 |
| 581-42-0 | 2,6-dimethylnaphthalene | 35.74 | 1406.5 | 70 |
| 112-54-9 | Dodecanal | 35.87 | 1409.4 | 96 |
| 475-20-7 | Longifolene | 36.03 | 1413.4 | 75 |
| 11028-42-5 | Cedrene | 36.32 | 1420.2 | 73 |
| 575-43-9 | 1,6-dimethylnaphthalene | 36.50 | 1424.5 | 90 |
| 575-37-1 | 1,7-dimethylnaphthalene | 36.54 | 1425.3 | 87 |
| 575-41-7 | 1,3-dimethylnaphthalene | 36.54 | 1425.4 | 82 |
| 101-81-5 | Diphenylmethane | 36.83 | 1432.1 | 74 |
| 131-11-3 | Dimethyl phthalate | 37.45 | 1446.8 | 92 |
| 112-53-8 | Dodecan-1-ol | 38.63 | 1475 | 98 |
| 473-13-2 | Alpha-selinene | 39.33 | 1491.6 | 89 |
| 17066-67-0 | Beta-selinene | 39.35 | 1492 | 88 |
| 13360-61-7 | 1-pentadecene | 39.36 | 1492.2 | 72 |
| 4630-07-3 | Valencene | 39.67 | 1499.6 | 72 |
| 629-62-9 | Pentadecane | 39.70 | 1500.2 | 95 |
| 96-76-4 | 2,4-di-tert-butylphenol | 39.88 | 1504.7 | 88 |
| 5875-45-6 | 2,5-di-tert-butylphenol | 39.92 | 1505.8 | 72 |
| 10486-19-8 | Tridecanal | 40.13 | 1511 | 87 |
| 39029-41-9 | Gamma-cadinene | 40.32 | 1515.7 | 77 |
| 132-64-9 | Dibenzofuran | 40.45 | 1518.9 | 83 |
| 483-76-1 | Delta-cadinene | 40.46 | 1519.3 | 81 |
| 21391-99-1 | Alpha-calacorene | 41.43 | 1543.6 | 86 |
| 4621-36-7 | 3-phenyldecane | 42.17 | 1561.9 | 84 |
| 134-62-3 | N,n-diethyl-m-toluamide | 42.59 | 1572.5 | 76 |
| 2050-08-0 | Amyl salicylate | 42.65 | 1574.1 | 73 |
| 84-66-2 | Diethyl phthalate | 43.12 | 1585.6 | 94 |
| 544-76-3 | Hexadecane | 43.70 | 1600.3 | 97 |
| 629-73-2 | 1-hexadecene | 43.73 | 1600.9 | 76 |
| 10233-13-3 | Isopropyl laurate | 44.65 | 1625.3 | 89 |
| 4537-14-8 | 6-phenylundecane | 44.72 | 1627.1 | 82 |
| 4537-15-9 | 5-phenylundecane | 44.86 | 1630.8 | 72 |
| 473-16-5 | Alpha-eudesmol | 45.46 | 1646.8 | 84 |
| 2156-97-0 | Lauryl acrylate | 47.20 | 1692.7 | 79 |
| 6765-39-5 | 1-heptadecene | 47.47 | 1699.7 | 86 |
| 629-78-7 | Heptadecane | 47.50 | 1700.5 | 96 |
| 24157-81-1 | 2,6-diisopropyl-naphthalene | 48.23 | 1720.8 | 76 |
| 2719-62-2 | 6-phenyldodecane | 48.41 | 1725.6 | 75 |
| 593-45-3 | Octadecane | 51.12 | 1800.7 | 96 |
| 118-60-5 | Octyl salicylate | 51.18 | 1802.3 | 85 |
| 638-36-8 | Phytane | 51.34 | 1806.8 | 79 |
| 110-27-0 | Isopropyl myristate | 51.92 | 1823.8 | 83 |
| 504-96-1 | Neophytadiene | 52.38 | 1837.2 | 72 |
| 1222-05-5 | Musk gx | 52.58 | 1842.9 | 84 |
| 629-92-5 | Nonadecane | 54.64 | 1903.1 | 71 |
| 107-06-2 | 1,2-dichloroethane | 4.61 | 649.6 | 92 |

**Table S 2:** ANOVA results for differences in the summed relative amount of each compound class identified between sampling sites on 27^th^ November, 2024.

| term | df | sumsq | meansq | statistic | p.value | Type_7 |
| --- | --- | --- | --- | --- | --- | --- |
| Location Name | 5 | 0.02 | 0.00 | 1.413450768 | 0.272 | Aliphatics |
| Location Name | 5 | 0.14 | 0.03 | 42.28108348 | <0.001 | Aromatics |
| Location Name | 5 | 0.10 | 0.02 | 56.94394467 | <0.001 | Monoterpenes |
| Location Name | 5 | 0.00 | 0.00 | 29.87459907 | <0.001 | Others (Esters, N, S, Halo) |
| Location Name | 5 | 0.00 | 0.00 | 6.986857086 | 0.001 | Sesquiterpenes |
| Location Name | 5 | 0.00 | 0.00 | 4.111920384 | 0.014 | Terpenoids |

**Table S 3:**
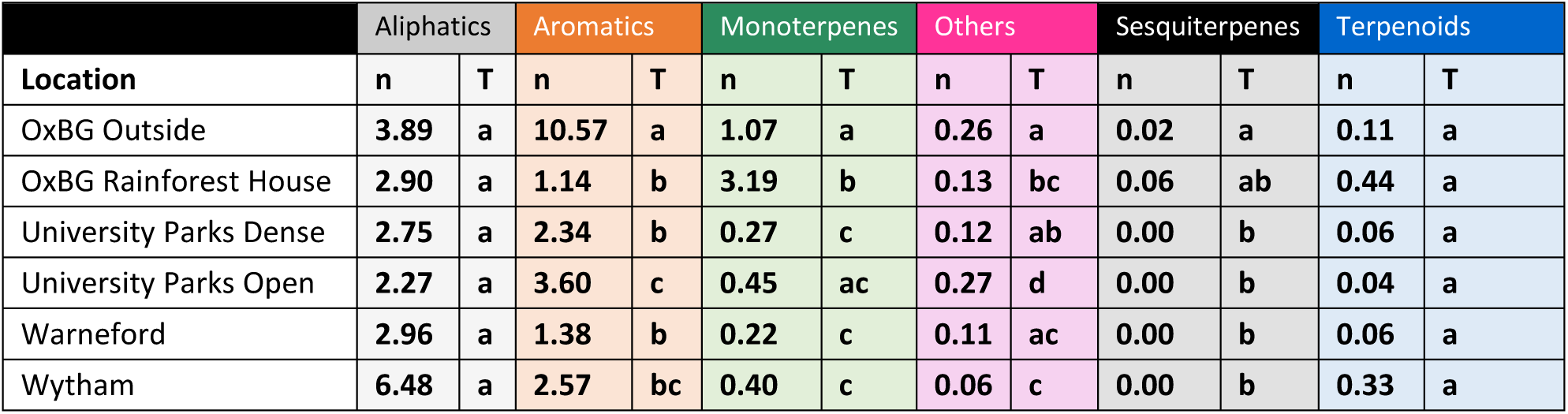
Summed relative amount of compounds identified within each compound class between sampling sites on 27^th^ November, 2024. Letters indicate groups differences based on Tukey post-hoc tests (p<0.05).

|  | Aliphatics |  | Aromatics |  | Monoterpenes |  | Others |  | Sesquiterpenes |  | Terpenoids |  |
| --- | --- | --- | --- | --- | --- | --- | --- | --- | --- | --- | --- | --- |
| Location | n | T | n | T | n | T | n | T | n | T | n | T |
| OxBG Outside | 3.89 | a | 10.57 | a | 1.07 | a | 0.26 | a | 0.02 | a | 0.11 | a |
| OxBG Rainforest House | 2.90 | a | 1.14 | b | 3.19 | b | 0.13 | bc | 0.06 | ab | 0.44 | a |
| University Parks Dense | 2.75 | a | 2.34 | b | 0.27 | c | 0.12 | ab | 0.00 | b | 0.06 | a |
| University Parks Open | 2.27 | a | 3.60 | c | 0.45 | ac | 0.27 | d | 0.00 | b | 0.04 | a |
| Warneford | 2.96 | a | 1.38 | b | 0.22 | c | 0.11 | ac | 0.00 | b | 0.06 | a |
| Wytham | 6.48 | a | 2.57 | bc | 0.40 | c | 0.06 | c | 0.00 | b | 0.33 | a |

**Table S 4:** ANOVA results for differences in the number of compounds identified within each compound class between sampling sites on 27^th^ November, 2024.

| term | df | sumsq | meansq | statistic | p.value | Type_7 |
| --- | --- | --- | --- | --- | --- | --- |
| Location Name | 5 | 364.54 | 72.91 | 2.435749877 | 0.080 | Aliphatics |
| Location Name | 5 | 331.04 | 66.21 | 4.607413753 | 0.009 | Aromatics |
| Location Name | 5 | 109.92 | 21.98 | 6.503543914 | 0.002 | Monoterpenes |
| Location Name | 5 | 69.17 | 13.83 | 3.124705882 | 0.037 | Others (Esters, N, S, Halo) |
| Location Name | 5 | 19.50 | 6.50 | 7.703703704 | 0.010 | Sesquiterpenes |
| Location Name | 5 | 167.61 | 33.52 | 20.89634002 | 0.000 | Terpenoids |

**Table S 5:** Summed number of compounds identified within each compound class between sampling sites on 27^th^ November, 2024. Letters indicate groups differences based on Tukey post-hoc tests (p<0.05).

| Location | Aliphatics |  | Aromatics |  | Monoterpenes |  | Others |  | Sesquiterpenes |  | Terpenoids |  |
| --- | --- | --- | --- | --- | --- | --- | --- | --- | --- | --- | --- | --- |
|  | n | T | n | T | n | T | n | T | n | T | n | T |
| OxBG Outside | n | Tuk | n | Tuk | n | Tuk | n | Tuk | n | Tuk | n | Tuk |
| OxBG Rainforest House | 65 | a | 41 | b | 13 | ab | 16 | b | 2 | a | 8 | b |
| University Parks Dense | 67 | a | 34 | ab | 15 | b | 15 | ab | 10 | b | 13 | c |
| University Parks Open | 60 | a | 29 | a | 9 | a | 9 | ab | 2 | a | 6 | ab |
| Warneford | 51 | a | 25 | a | 9 | a | 10 | ab | 1 | a | 4 | a |
| Wytham | 59 | a | 36 | ab | 8 | a | 11 | ab | 1 | a | 5 | ab |

**Table S 6:**
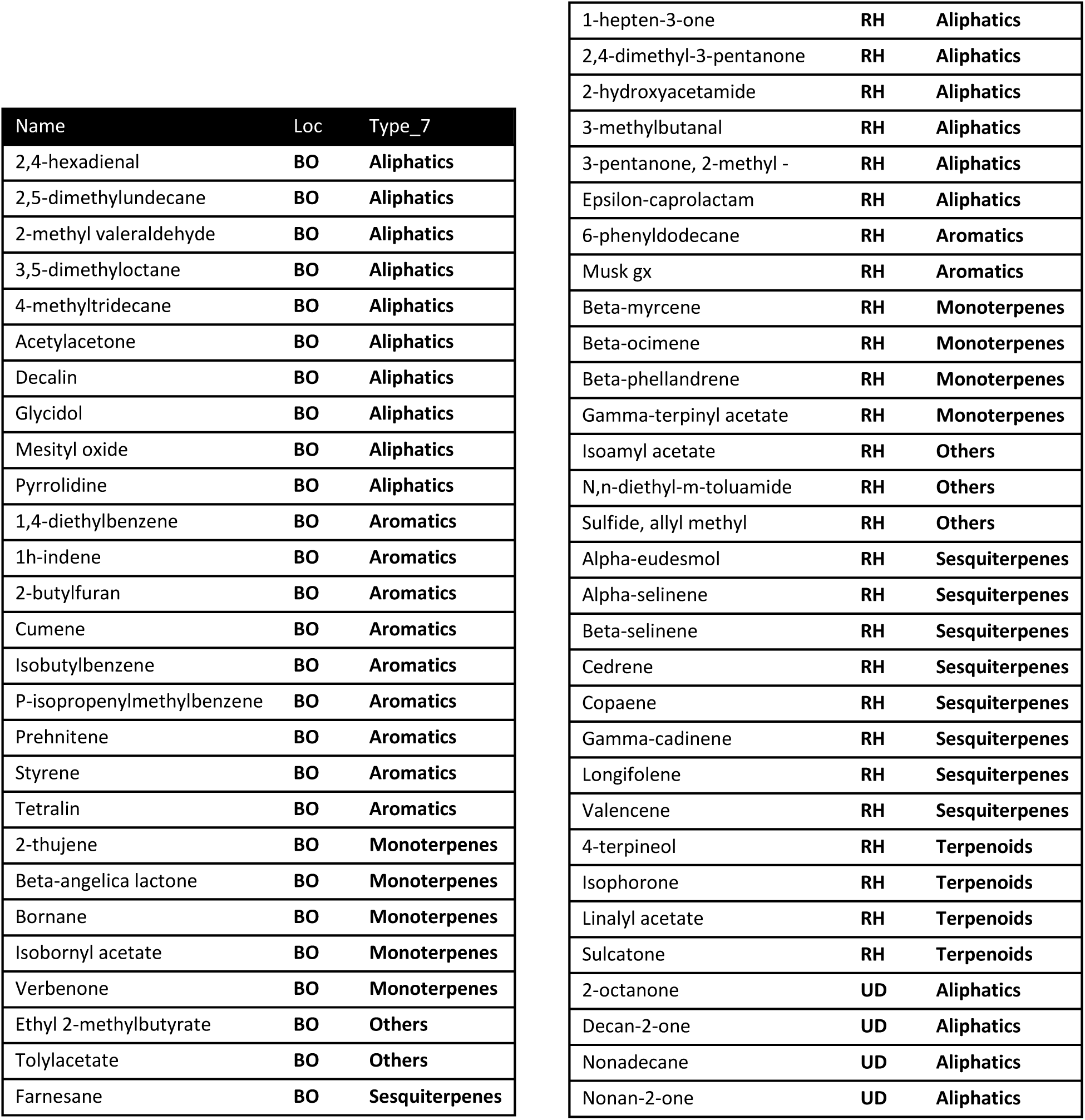
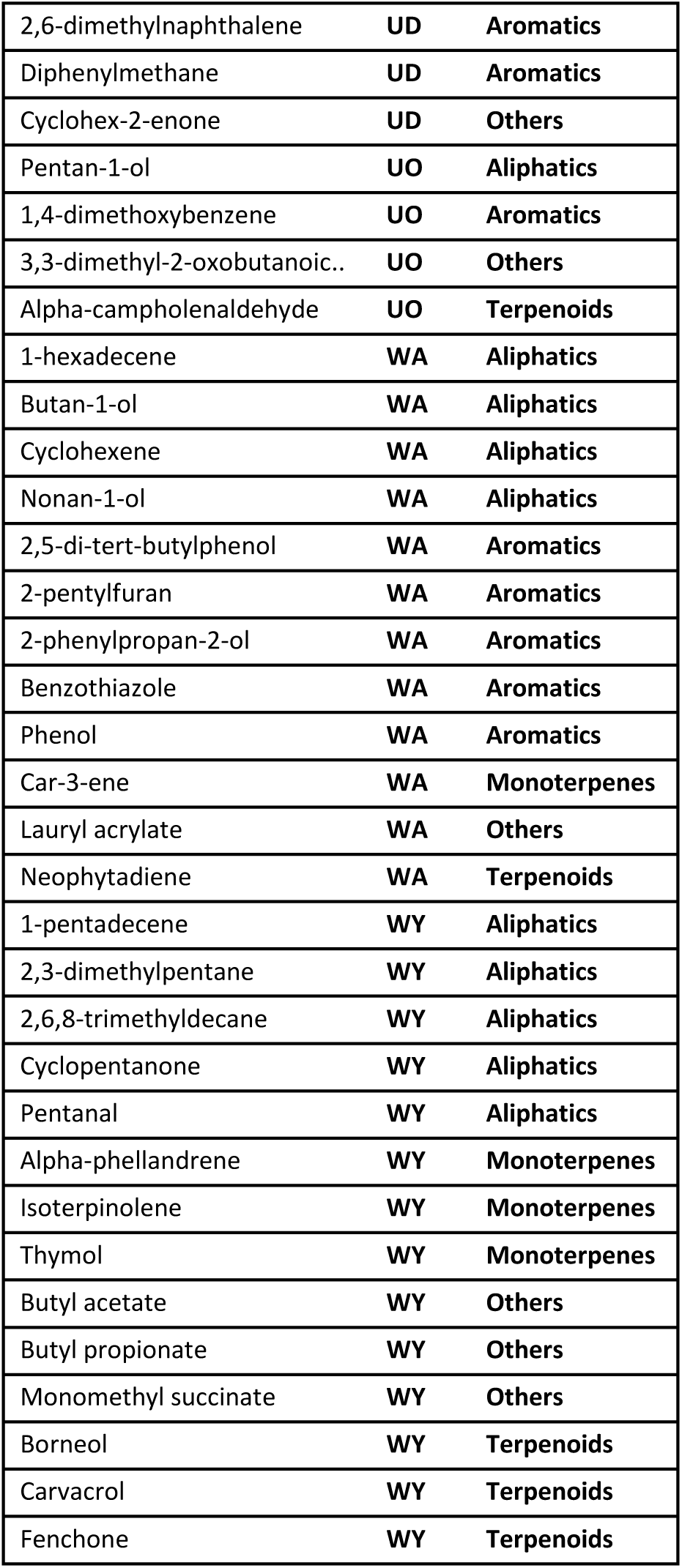
All compounds identified only at a single site during the 27 November 2024 sampling.

**Table S 7:** Top-ranking compounds from PLS-DA analyses with explicit directionality. VIP scores (> ranked top 10) identify compounds contributing most strongly to group separation. Group-wise mean abundances are provided to support directional interpretation.

| Comparison | Direction | Compound | VIP | Loading | Mean_Group1 | Mean_Group2 |
| --- | --- | --- | --- | --- | --- | --- |
| WAH/WYH/UDH/RHH v BOH/UOH | > in BOH/UOH | P-xylene (Aromatics) | 2.296 | -0.148 | 0.186 | 0.367 |
| WAH/WYH/UDH/RHH v BOH/UOH | > in BOH/UOH | Ethylbenzene (Aromatics) | 2.268 | -0.146 | 0.108 | 0.209 |
| WAH/WYH/UDH/RHH v BOH/UOH | > in BOH/UOH | Isooctane (Aliphatics) | 2.206 | -0.142 | 0.035 | 0.133 |
| WAH/WYH/UDH/RHH v BOH/UOH | > in BOH/UOH | 2,3,4-trimethylpentane (Aliphatics) | 2.150 | -0.138 | 0.032 | 0.087 |
| WAH/WYH/UDH/RHH v BOH/UOH | > in BOH/UOH | Toluene (Aromatics) | 2.138 | -0.137 | 0.239 | 0.424 |
| WAH/WYH/UDH/RHH v BOH/UOH | > in BOH/UOH | 3-methylhexane (Aliphatics) | 2.121 | -0.136 | 0.064 | 0.136 |
| WAH/WYH/UDH/RHH v BOH/UOH | > in BOH/UOH | Methylcyclohexane (Aliphatics) | 2.065 | -0.133 | 0.065 | 0.131 |
| WAH/WYH/UDH/RHH v BOH/UOH | > in WAH/WYH/UDH/RHH | 2-dodecene (Aliphatics) | 2.060 | 0.132 | 0.198 | 0.000 |
| WAH/WYH/UDH/RHH v BOH/UOH | > in BOH/UOH | 1-ethyl-2-methylbenzene (Aromatics) | 2.014 | -0.129 | 0.027 | 0.110 |
| WAH/WYH/UDH/RHH v BOH/UOH | > in BOH/UOH | 3-methylheptane (Aliphatics) | 2.005 | -0.129 | 0.041 | 0.074 |
| WAH/WYH/UDH v RHH | > in RHH | Beta-ocimene (Monoterpenes) | 2.285 | -0.147 | 0.000 | 0.072 |
| WAH/WYH/UDH v RHH | > in RHH | Dihydromyrcenol (Terpenoids) | 2.281 | -0.147 | 0.000 | 0.105 |
| WAH/WYH/UDH v RHH | > in RHH | Sabinene (Monoterpenes) | 2.273 | -0.146 | 0.005 | 0.374 |
| WAH/WYH/UDH v RHH | > in RHH | Beta-pinene (Monoterpenes) | 2.244 | -0.144 | 0.033 | 0.287 |
| WAH/WYH/UDH v RHH | > in RHH | Pinocarvone (Terpenoids) | 2.232 | -0.143 | 0.000 | 0.043 |
| WAH/WYH/UDH v RHH | > in RHH | Hydrindane (Aliphatics) | 2.216 | -0.142 | 0.000 | 0.028 |
| WAH/WYH/UDH v RHH | > in RHH | Alpha-pinene (Monoterpenes) | 2.196 | -0.141 | 0.104 | 0.376 |
| WAH/WYH/UDH v RHH | > in RHH | Copaene (Sesquiterpenes) | 2.195 | -0.141 | 0.000 | 0.048 |
| WAH/WYH/UDH v RHH | > in RHH | 4-terpineol (Terpenoids) | 2.195 | -0.141 | 0.000 | 0.034 |
| WAH/WYH/UDH v RHH | > in RHH | Beta-phellandrene (Monoterpenes) | 2.169 | -0.139 | 0.000 | 0.045 |
| BOH v UOH/UDH | > in BOH | 2-thujene (Monoterpenes) | 2.300 | -0.148 | 0.055 | 0.000 |
| BOH v UOH/UDH | > in BOH | Prehnitene (Aromatics) | 2.288 | -0.147 | 0.035 | 0.000 |
| BOH v UOH/UDH | > in BOH | Farnesane (Sesquiterpenes) | 2.284 | -0.147 | 0.035 | 0.000 |
| BOH v UOH/UDH | > in BOH | P-isopropenylmethylbenzene (Aromatics) | 2.274 | -0.146 | 0.020 | 0.000 |
| BOH v UOH/UDH | > in BOH | Bromodichloromethane (Others (Esters, N...)) | 2.268 | -0.146 | 0.013 | 0.000 |
| BOH v UOH/UDH | > in BOH | 1,4-diethylbenzene (Aromatics) | 2.259 | -0.145 | 0.045 | 0.000 |
| BOH v UOH/UDH | > in UOH/UDH | Tricyclene (Monoterpenes) | 2.251 | 0.145 | 0.000 | 0.076 |
| BOH v UOH/UDH | > in BOH | 2,2,4,6,6-pentamethylheptane (Aliphatics) | 2.201 | -0.142 | 0.099 | 0.062 |
| BOH v UOH/UDH | > in BOH | Bornyl acetate (Terpenoids) | 2.183 | -0.140 | 0.017 | 0.000 |
| BOH v UOH/UDH | > in BOH | Propylbenzene (Aromatics) | 2.146 | -0.138 | 0.093 | 0.009 |

