## Supplementary Information for "A walk in the park – identifying healthy greenspaces using scents" (from another version of this preprint: version 1, 2025-08-29)

### A walk in the park – identifying healthy biomes using scent snapshots

#### 1 Supplementary Information

**Table S 1:** A full list of the compounds detected in this study including retention time, and compound class. List is sorted by retention time (RT). Others represent Esters, Nitrogen containing compounds, Sulphur containing compounds, and Halogenated compounds)

| CAS | Name | RT | Class |
|---|---|---|---|
| 78-84-2 | Isobutyraldehyde | 3.3837 | Aliphatics |
| 142-29-0 | Cyclopentene | 3.4257 | Aliphatics |
| 96-14-0 | 3-methylpentane | 3.8344 | Aliphatics |
| 431-03-8 | Butane-2,3-dione | 3.86 | Aliphatics |
| 78-93-3 | Butan-2-one | 3.9178 | Aliphatics |
| 123-72-8 | Butanal | 3.921 | Aliphatics |
| 110-54-3 | Hexane | 3.941 | Aliphatics |
| 930-27-8 | 3-methylfuran | 4.0033 | Aromatics |
| 534-22-5 | 2-methylfuran | 4.0107 | Aromatics |
| 115-18-4 | 3-hydroxy-3-methylbut-1-ene | 4.0746 | Aliphatics |
| 141-78-6 | Ethyl acetate | 4.0788 | Others |
| 67-66-3 | Chloroform | 4.2538 | Others |
| 109-99-9 | Oxolane | 4.2811 | Aliphatics |
| 96-37-7 | Methylcyclopentane | 4.3198 | Aliphatics |
| 107-06-2 | 1,2-dichloroethane | 4.6231 | Others |
| 563-80-4 | 3-methylbutan-2-one | 4.6489 | Aliphatics |
| 71-43-2 | Benzene | 4.7352 | Aromatics |
| 56-23-5 | Tetrachloromethane | 4.7685 | Others |
| 71-36-3 | Butan-1-ol | 4.7928 | Aliphatics |
| 110-82-7 | Cyclohexane | 4.7997 | Aliphatics |
| 589-34-4 | 3-methylhexane | 4.8724 | Aliphatics |
| 617-78-7 | 3-ethylpentane | 5.0631 | Aliphatics |
| 107-87-9 | Pentan-2-one | 5.0788 | Aliphatics |
| 540-84-1 | Isooctane | 5.0984 | Aliphatics |
| 142-82-5 | Heptane | 5.2872 | Aliphatics |
| 79-01-6 | Trichloroethene | 5.3766 | Others |
| 80-62-6 | Methyl methacrylate | 5.501 | Others |
| 109-60-4 | Propyl acetate | 5.5457 | Others |
| 625-86-5 | 2,5-dimethylfuran | 5.5939 | Aromatics |
| 3208-16-0 | 2-ethylfuran | 5.604 | Aromatics |
| 75-27-4 | Bromodichloromethane | 5.6272 | Others |
| 18936-17-9 | 2-methylbutanenitrile | 5.8016 | Aliphatics |
| 108-87-2 | Methylcyclohexane | 5.9185 | Aliphatics |
| 589-43-5 | 2,4-dimethylhexane | 5.9796 | Aliphatics |
| 123-51-3 | Isoamylol | 6.0643 | Aliphatics |
| 137-32-6 | 2-methylbutan-1-ol | 6.0678 | Aliphatics |
| 108-10-1 | Methyl isobutyl ketone | 6.152 | Aliphatics |
| 624-92-0 | Dimethyl disulfide | 6.3619 | Others |
| 565-69-5 | 3-pentanone, 2-methyl - | 6.4284 | Aliphatics |
| 565-75-3 | 2,3,4-trimethylpentane | 6.4487 | Aliphatics |
| 563-16-6 | 3,3-dimethylhexane | 6.6135 | Aliphatics |
| 560-21-4 | 2,3,3-trimethylpentane | 6.6313 | Aliphatics |
| 589-53-7 | 4-methylheptane | 6.8642 | Aliphatics |
| 108-88-3 | Toluene | 6.9307 | Aromatics |
| 589-81-1 | 3-methylheptane | 7.0475 | Aliphatics |
| 111-66-0 | 1-octene | 7.5919 | Aliphatics |
| 3848-24-6 | Hexane-2,3-dione | 7.6407 | Aliphatics |
| 111-65-9 | Octane | 7.8792 | Aliphatics |
| 66-25-1 | Hexanal | 7.9063 | Aliphatics |
| 589-90-2 | 1,4-dimethylcyclohexane | 7.9708 | Aliphatics |
| 124-48-1 | Chlorodibromomethane | 8.023 | Others |
| 127-18-4 | Tetrachloroethene | 8.1201 | Others |
| 106-36-5 | Propyl propionate | 8.1825 | Others |
| 2213-23-2 | 2,4-dimethylheptane | 8.6053 | Aliphatics |
| 123-42-2 | Diacetone alcohol | 9.289 | Aliphatics |
| 19549-87-2 | 2,4-dimethyl-1-heptene | 9.3148 | Aliphatics |
| 108-90-7 | Chlorobenzene | 9.581 | Aromatics |
| 100-41-4 | Ethylbenzene | 10.1506 | Aromatics |
| 106-42-3 | P-xylene | 10.5131 | Aromatics |
| 123-92-2 | Isoamyl acetate | 10.7472 | Others |
| 106-35-4 | Heptan-3-one | 11.1233 | Aliphatics |
| 124-11-8 | 1-nonene | 11.2049 | Aliphatics |
| 110-43-0 | Heptan-2-one | 11.2844 | Aliphatics |
| 100-42-5 | Styrene | 11.4281 | Aromatics |
| 108-94-1 | Cyclohexanone | 11.5463 | Aliphatics |
| 111-84-2 | Nonane | 11.7724 | Aliphatics |
| 111-71-7 | Heptanal | 11.8431 | Aliphatics |
| 111-76-2 | 2-butoxyethanol | 11.9618 | Aliphatics |
| 15869-93-9 | 3,5-dimethyloctane | 12.7417 | Aliphatics |
| 508-32-7 | Tricyclene | 12.8312 | Monoterpenes |
| 98-82-8 | Cumene | 12.8383 | Aromatics |
| 4110-44-5 | 3,3-dimethyloctane | 13.2931 | Aliphatics |
| 7785-26-4 | Alpha-pinene | 13.3298 | Monoterpenes |
| 79-92-5 | Camphene | 14.1187 | Monoterpenes |
| 103-65-1 | Propylbenzene | 14.2346 | Aromatics |
| 123-05-7 | 2-ethylhexanal | 14.2447 | Aliphatics |
| 620-14-4 | 3-ethyltoluene | 14.5823 | Aromatics |
| 100-52-7 | Benzaldehyde | 14.7628 | Aromatics |
| 108-67-8 | 1,3,5-trimethylbenzene | 14.9842 | Aromatics |
| 28634-89-1 | 2-thujene | 15.1858 | Monoterpenes |
| 3387-41-5 | Sabinene | 15.1918 | Monoterpenes |
| 99-82-1 | P-menthane | 15.2135 | Monoterpenes |
| 464-15-3 | Bornane | 15.314 | Monoterpenes |
| 611-14-3 | 1-ethyl-2-methylbenzene | 15.416 | Aromatics |
| 18172-67-3 | Beta-pinene | 15.4411 | Monoterpenes |
| 110-93-0 | Sulcatone | 15.703 | Terpenoids |
| 872-05-9 | 1-decene | 15.7323 | Aliphatics |

| | | | |
|---|---|---|---|
| 100-47-0 | Benzonitrile | 15.7966 | Aromatics |
| 123-35-3 | Beta-myrcene | 15.9591 | Monoterpenes |
| 13475-82-6 | 2,2,4,6,6-pentamethylheptane | 15.9835 | Aliphatics |
| 4551-51-3 | Hydrindane | 16.1091 | Aliphatics |
| 95-63-6 | 1,2,4-trimethylbenzene | 16.1804 | Aromatics |
| 288-88-0 | 1h-1,2,4-triazole | 16.3004 | Aromatics |
| 124-18-5 | Decane | 16.4858 | Aliphatics |
| 124-13-0 | Octanal | 16.6393 | Aliphatics |
| 13466-78-9 | 3-carene | 16.925 | Monoterpenes |
| 111-90-0 | Diethylene glycol monoethyl ether | 17.0248 | Aliphatics |
| 99-86-5 | Alpha-terpinene | 17.3454 | Monoterpenes |
| 526-73-8 | 1,2,3-trimethylbenzene | 17.4966 | Aromatics |
| 99-87-6 | P-cymene | 17.7153 | Monoterpenes |
| 527-84-4 | O-cymene | 17.7252 | Aromatics |
| 535-77-3 | M-cymene | 17.7382 | Aromatics |
| 104-76-7 | 2-ethylhexan-1-ol | 17.8599 | Aliphatics |
| 5989-27-5 | Limonene | 17.9441 | Monoterpenes |
| 555-10-2 | Beta-phellandrene | 18.0317 | Monoterpenes |
| 470-82-6 | 1,8-cineole | 18.1177 | Terpenoids |
| 496-11-7 | Indane | 18.1741 | Aromatics |
| 13877-91-3 | Beta-ocimene | 18.2467 | Monoterpenes |
| 95-13-6 | 1h-indene | 18.6427 | Aromatics |
| 122-78-1 | Phenylacetaldehyde | 18.7082 | Aromatics |
| 105-05-5 | 1,4-diethylbenzene | 18.8056 | Aromatics |
| 99-85-4 | Gamma-terpinene | 19.375 | Monoterpenes |
| 493-02-7 | Decahydronaphthalene | 19.4565 | Aliphatics |
| 98-86-2 | Acetophenone | 19.7822 | Aromatics |
| 18479-58-8 | Dihydromyrcenol | 20.0559 | Terpenoids |
| 1195-32-0 | P-isopropenylmethyl benzene | 20.4872 | Aromatics |
| 617-94-7 | 2-phenylpropan-2-ol | 20.7675 | Aromatics |
| 78-69-3 | Linalool | 21.4178 | Terpenoids |
| 1120-21-4 | Undecane | 21.4477 | Aliphatics |
| 124-19-6 | Nonanal | 21.682 | Aliphatics |
| 546-80-5 | Alpha-thujone | 21.8054 | Terpenoids |
| 95-93-2 | Durene | 22.4144 | Aromatics |
| 76-22-2 | Camphor | 23.8474 | Terpenoids |
| 488-23-3 | Prehnitene | 23.9119 | Monoterpenes |
| 89-80-5 | Menthone | 24.2345 | Terpenoids |
| 119-64-2 | Tetralin | 24.4536 | Aromatics |
| 140-39-6 | Tolylacetate | 24.5419 | Others |
| 30460-92-5 | Pinocarvone | 24.5623 | Terpenoids |
| 2216-51-5 | Menthol | 25.3144 | Terpenoids |
| 562-74-3 | 4-terpineol | 25.4509 | Terpenoids |
| 91-20-3 | Naphthalene | 25.6484 | Aromatics |
| 119-36-8 | Methyl salicylate | 25.9934 | Aromatics |
| 112-40-3 | Dodecane | 26.3416 | Aliphatics |
| 112-31-2 | Decanal | 26.6995 | Aliphatics |
| 1196-01-6 | Verbenone | 26.7273 | Monoterpenes |
| 7206-26-0 | 2-dodecene | 26.8779 | Aliphatics |
| 17301-22-3 | 2,5-dimethylundecane | 26.9371 | Aliphatics |
| 6485-40-1 | Carvone | 28.4412 | Terpenoids |
| 115-95-7 | Linalyl acetate | 28.662 | Terpenoids |
| 112-05-0 | Nonanoic acid | 29.4373 | Aliphatics |
| 31295-56-4 | 2,6,11-trimethyldodecane | 29.8235 | Aliphatics |
| 20347-65-3 | Bornyl acetate | 30.4542 | Terpenoids |
| 91-57-6 | 2-methylnaphthalene | 30.9567 | Aromatics |
| 629-50-5 | Tridecane | 31.0369 | Aliphatics |
| 112-44-7 | Undecanal | 31.376 | Aliphatics |
| 90-12-0 | 1-methylnaphthalene | 31.5739 | Aromatics |
| 26730-12-1 | 4-methyltridecane | 33.8909 | Aliphatics |
| 3891-98-3 | Farnesane | 34.3715 | Sesquiterpenes |
| 3856-25-5 | Copaene | 34.5267 | Sesquiterpenes |
| 1120-36-1 | 1-tetradecene | 35.1371 | Aliphatics |
| 80-46-6 | P-t-amylphenol | 35.1785 | Aromatics |
| 629-59-4 | Tetradecane | 35.474 | Aliphatics |
| 112-54-9 | Dodecanal | 35.8582 | Aliphatics |
| 575-43-9 | 1,6-dimethylnaphthalene | 36.5648 | Aromatics |
| 131-11-3 | Dimethyl phthalate | 37.46 | Others |
| 112-53-8 | Dodecan-1-ol | 38.681 | Aliphatics |
| 629-62-9 | Pentadecane | 39.7013 | Aliphatics |
| 96-76-4 | 2,4-di-tert-butylphenol | 39.8783 | Aromatics |
| 10486-19-8 | Tridecanal | 40.1348 | Aliphatics |
| 483-76-1 | Delta-cadinene | 40.4941 | Sesquiterpenes |
| 21391-99-1 | Alpha-calacorene | 41.4255 | Sesquiterpenes |
| 4621-36-7 | 3-phenyldecane | 42.1656 | Aromatics |
| 84-66-2 | Diethyl phthalate | 43.107 | Aromatics |
| 544-76-3 | Hexadecane | 43.7033 | Aliphatics |
| 10233-13-3 | Isopropyl laurate | 44.6496 | Others |
| 4537-14-8 | 6-phenylundecane | 44.7236 | Aromatics |
| 6765-39-5 | 1-heptadecene | 47.4592 | Aliphatics |
| 629-78-7 | Heptadecane | 47.5002 | Aliphatics |
| 24157-81-1 | 2,6-diisopropylnaphthalene | 48.2415 | Aromatics |
| 593-45-3 | Octadecane | 51.1247 | Aliphatics |
| 118-60-5 | Octyl salicylate | 51.187 | Aromatics |
| 638-36-8 | Phytane | 51.3357 | Terpenoids |
| 110-27-0 | Isopropyl myristate | 51.9204 | Others |
| 1222-05-5 | Musk gx | 52.5787 | Aromatics |
| 112-95-8 | Icosane | 57.9363 | Aliphatics |



**Table S 2:** The top 40 compounds contributing to the differences between sampling sites from the PCA.

Ethylbenzene, P-xylene, Linalool, Menthone, Gamma-terpinene, Toluene, Sabinene, Beta-ocimene, Copaene, Ethyl acetate, 1,2,4-trimethylbenzene, Pinocarvone, Pentadecane, Beta-pinene, Decahydronaphthalene, 4,5-dimethyl-1-hexene, Dodecan-1-ol, 1,4-dimethylcyclohexane, 4-terpineol, Benzaldehyde, Acetophenone, Dodecanal, Alpha-pinene, 1-nonene, 1,3,5-trimethylbenzene, Heptane, Heptadecane, Hexane, 2,4-dimethylheptane, 2-ethylhexan-1-ol, Octadecane, 2,3,4-trimethylpentane, 3-ethyltoluene, 2,3-dimethylpentane, 2,2,4,6,6-pentamethylheptane, 3,3-dimethyloctane, Menthol, 2-chlorobutane, Butan-2-one, Methylcyclohexane.

**Table S 3:** Vegetation surveys carried out at six sampling sites using quadrats. Botanical Gardens Outdoors (BO), OxBG Rainforest Glasshouse (RH), University Parks Dense (UD), #University Parks Open (UO), Warneford Meadow (WA), Wytham Woods (WY).

| | |
|---|---|
| BO | *Cordyline australis* |
| BO | *Galanthus galatea* |
| BO | *Hamamelis 'Orange Peel'* |
| BO | *Junpiperus chinensis* |
| BO | *Laurus azorica* |
| BO | *Liriodendron chinense* |
| BO | *Magnolia doltsopa* |
| BO | *Juniperus chinensis* |
| BO | *Allium flavum* |
| BO | *Allium karataviense* |
| BO | *Allium oreophilum* |
| BO | *Allium rosenbachianum* |
| BO | *Allium siculum* |
| BO | *Allium sphaerocephalum* |
| BO | *Aristolochia tomentosa* |
| BO | *Aster divaricatus* |
| BO | *Bletilla striata 'Albo-striata'* |
| BO | *Camassia cusickii* |
| BO | *Chamaerops humilis* |
| BO | *Cortaderia selloana* |
| BO | *Crocus sieberi 'Firefly'* |
| BO | *Crocus speciosus* |
| BO | *Fargesia scabrida* |
| BO | *Galium odoratum* |
| BO | *Geranium sylvaticum 'Mayflower'* |
| BO | *Helleborus orientalis 'Ballardâ€™s hybrids'* |
| BO | *Iris danfordiae* |
| BO | *Kniphofia citrina* |
| BO | *Lilium martagon 'Album'* |
| BO | *Lilium regale* |
| BO | *Lilium regale* |
| BO | *Narcissus Ã— incomparabilis* |
| BO | *Narcissus lobularis* |
| BO | *Narcissus poeticus var. recurvus* |
| BO | *Nassella trichotoma* |
| BO | *Sauromatum venustum* |
| BO | *Scilla bifolia* |
| BO | *Scilla mischtschenkoana* |
| BO | *Scilla peruviana* |
| BO | *Sternbergia lutea* |
| BO | *Tulipa kolpakowskiana* |
| BO | *Tulipa turkestanica* |
| BO | *Yucca gloriosa 'Variegata'* |
| BO | *Zantedeschia albomaculata* |
| RH | *Acalypha chamaedrifolia* |
| RH | *Acalypha wilkesiana* |
| RH | *Adiantum macrophyllum* |
| RH | *Adonidia merrillii* |
| RH | *Aechmea aquilega var. aquilega* |
| RH | *Aechmea bracteata* |
| RH | *Aechmea bromeliifolia* |
| RH | *Aechmea chantinii* |
| RH | *Aechmea fosteriana* |
| RH | *Aechmea mexicana* |
| RH | *Aechmea pineliana* |
| RH | *Aechmea purpureorosea* |
| RH | *Aechmea racinae* |
| RH | *Aechmea recurvata* |
| RH | *Aeschynanthus gracilis* |
| RH | *Aframomum melegueta* |
| RH | *Aglaia elliptica subsp. elliptica* |
| RH | *Aglaia odorata* |
| RH | *Aglaonema* |
| RH | *Alocasia macrorrhizos* |
| RH | *Alocasia odora* |
| RH | *Alpinia calcarata* |
| RH | *Alpinia haenkei* |
| RH | *Alpinia purpurata* |
| RH | *Alpinia zerumbet* |
| RH | *Amorphophallus henryi* |
| RH | *Amorphophallus konjac* |
| RH | *Amorphophallus konjac* |
| RH | *Amorphophallus krausei* |
| RH | *Amorphophallus titanum* |
| RH | *Amydrium zippelianum* |
| RH | *Ananas comosus var. bracteatus* |
| RH | *Angraecum eburneum* |
| RH | *Annona senegalensis* |
| RH | *Anthorrhiza echinella* |
| RH | *Anthurium* |
| RH | *Anthurium* |
| RH | *Anthurium* |
| RH | *Anthurium gracile* |
| RH | *Anthurium obtusum subsp. obtusum* |
| RH | *Antidesma montanum var. montanum* |
| RH | *Ardisia crenata* |
| RH | *Arenga caudata* |
| RH | *Aristolochia arborea* |
| RH | *Aristolochia liukiuensis* |
| RH | *Aristolochia salvadorensis* |
| RH | *Aristolochia zollingeriana* |
| RH | *Artabotrys hexapetalus* |
| RH | *Asplenium nidus* |
| RH | *Austrobaileya scandens* |
| RH | *Averrhoa carambola* |
| RH | *Barleria cristata* |
| RH | *Barleria obtusa* |
| RH | *Barleria oenotheroides* |
| RH | *Barleria prionitis* |
| RH | *Beaumontia grandiflora* |
| RH | *Begonia arborescens* |
| RH | *Begonia 'Compte de Lesseps'* |
| RH | *Begonia dietrichiana* |
| RH | *Begonia 'Fireworks'* |
| RH | *Begonia listada* |
| RH | *Begonia manicata 'Crispa'* |
| RH | *Begonia masoniana* |
| RH | *Begonia oxyphylla* |
| RH | *Begonia reniformis* |
| RH | *Begonia sericoneura* |
| RH | *Begonia serratipetala* |
| RH | *Begonia solimutata* |
| RH | *Begonia 'Superba'* |
| RH | *Billbergia amoena* |
| RH | *Billbergia nutans* |
| RH | *Billbergia zebrina* |
| RH | *Billbergia zebrina* |
| RH | *Blechnum* |
| RH | *Blechnum neohollandicum* |
| RH | *Brexia madagascariensis* |
| RH | *Breynia disticha* |
| RH | *Brillantaisia lamium* |
| RH | *Bromelia karatas* |
| RH | *Burbidgea schizocheila* |
| RH | *Butia odorata* |
| RH | *Caladium bicolor* |
| RH | *Calathea* |
| RH | *Camptotheca acuminata* |
| RH | *Canistropsis billbergioides* |
| RH | *Canistropsis burchellii* |
| RH | *Canistropsis burchellii* |
| RH | *Canna × ehemannii* |
| RH | *Canna indica* |
| RH | *Carapichea ipecacuanha* |
| RH | *Carica papaya* |
| RH | *Carludovica palmata* |
| RH | *Carpentaria acuminata* |
| RH | *Catha edulis* |
| RH | *Catha edulis* |
| RH | *Catharanthus trichophyllus* |
| RH | *Ceratozamia kuesteriana* |
| RH | *Ceratozamia mexicana* |
| RH | *Ceratozamia robusta* |
| RH | *Chamaedorea stolonifera* |
| RH | *Chlorophytum madagascariense* |
| RH | *Chlorophytum sparsiflorum* |
| RH | *Cibotium regale* |
| RH | *Cinchona calisaya* |
| RH | *Cinnamomum verum* |
| RH | *Cissus discolor* |
| RH | *Citrus × aurantium* |
| RH | *Coffea arabica* |
| RH | *Coffea arabica 'Bourbon'* |

| | |
|---|---|
| RH | *Coffea benghalensis* |
| RH | *Coffea canephora* |
| RH | *Coffea liberica* |
| RH | *Coffea mauritiana* |
| RH | *Coffea stenophylla* |
| RH | *Colocasia esculenta* |
| RH | *Colocasia esculenta 'Maui Gold'* |
| RH | *Costus afer* |
| RH | *Costus curvibracteatus* |
| RH | *Costus pictus* |
| RH | *Crossandra flava* |
| RH | *Crossandra nilotica* |
| RH | *Cryosophila warscewiczii* |
| RH | *Cryptostegia grandiflora* |
| RH | *Ctenanthe lubbersiana* |
| RH | *Ctenanthe setosa* |
| RH | *Cubanola domingensis* |
| RH | *Curculigo capitulata* |
| RH | *Curcuma alismatifolia* |
| RH | *Curcuma longa* |
| RH | *Curcuma zedoaria* |
| RH | *Cycas rumphii* |
| RH | *Cymbopogon citratus* |
| RH | *Davallia trichomanoides* |
| RH | *Deherainia smaragdina* |
| RH | *Dichorisandra thyrsiflora* |
| RH | *Dicliptera raui* |
| RH | *Dieffenbachia seguine* |
| RH | *Dioon pectinatum* |
| RH | *Dioscorea* |
| RH | *Dorstenia arifolia* |
| RH | *Dorstenia contrajerva* |
| RH | *Drymophloeus litigiosus* |
| RH | *Drynaria meyeniana* |
| RH | *Dypsis decaryi* |
| RH | *Dypsis lutescens* |
| RH | *Elettaria cardamomum* |
| RH | *Elettaria ensal* |
| RH | *Encephalartos ferox* |
| RH | *Ensete ventricosum* |
| RH | *Epipremnum aureum* |
| RH | *Epipremnum pinnatum* |
| RH | *Eupomatia bennettii* |
| RH | *Eupomatia bennettii* |
| RH | *Ficus sagittifolia* |
| RH | *Fittonia gigantea* |
| RH | *Geogenanthus poeppigii* |
| RH | *Globba marantina* |
| RH | *Gnetum gnemon* |
| RH | *Gossypium arboreum* |
| RH | *Gossypium darwinii* |
| RH | *Gossypium raimondii* |
| RH | *Graptophyllum pictum* |
| RH | *Gravesia guttata* |
| RH | *Guzmania wittmackii* |
| RH | *Hedychium coronarium* |
| RH | *Hedychium greenii* |
| RH | *Hedychium hasseltii* |
| RH | *Hedychium muluense* |
| RH | *Heliconia rostrata* |
| RH | *Heliconia stricta 'Ali'* |
| RH | *Hemionitis palmata* |
| RH | *Hernandia nymphaeifolia* |
| RH | *Hevea brasiliensis* |
| RH | *Hibiscus acetosella* |
| RH | *Hibiscus sabdariffa* |
| RH | *Hibiscus schizopetalus* |
| RH | *Hibiscus taiwanensis* |
| RH | *Hilleria latifolia* |
| RH | *Howea belmoreana* |
| RH | *Howea forsteriana* |
| RH | *Hydnophytum formicarum* |
| RH | *Hydnophytum moseleyanum* |
| RH | *Hydnophytum radicans* |
| RH | *Hypoestes* |
| RH | *Hypolepis dicksonioides* |
| RH | *Ilex paraguariensis* |
| RH | *Impatiens niamniamensis* |
| RH | *Impatiens sodenii* |
| RH | *Ipomoea trifida* |
| RH | *Jacquinia arborea* |
| RH | *Justicia scheidweileri* |
| RH | *Justicia spicigera* |
| RH | *Leea guineensis* |
| RH | *Lepismium cruciforme* |
| RH | *Leucocasia gigantea* |
| RH | *Lippia dulcis* |
| RH | *Livistona saribus* |
| RH | *Lomariopsis tenuifolia* |
| RH | *Macadamia integrifolia* |
| RH | *Macrocentrum cristatum* |
| RH | *Malpighia glabra* |
| RH | *Manihot esculenta* |
| RH | *Manilkara zapota* |
| RH | *Maranta arundinacea* |
| RH | *Marsdenia formosana* |
| RH | *Microlepia speluncae* |
| RH | *Microsorum* |
| RH | *Mirabilis jalapa* |
| RH | *Monstera adansonii* |
| RH | *Morisonia paradoxa* |
| RH | *Mucuna macrocarpa* |
| RH | *Muehlenbeckia platyclada* |
| RH | *Murdannia loriformis* |
| RH | *Murraya paniculata* |
| RH | *Musa acuminata* |
| UD | *Taraxacum officinale* |
| UD | *Hedera helix* |
| UD | *Petroselinum crispum* |
| UD | *Trillium chloropetalum* |
| UD | *Iris foetidissima* |
| UD | *Trifolium repens* |
| UD | *Bellis perennis* |
| UD | *Geum urbanum* |
| UD | *Ranunculus repens* |
| UD | *Primula hendersonii* |
| UD | *Glechoma hederacea* |
| UD | *Viburnum rhytidophyllum* |
| UD | *Aesculus hippocastanum* |
| UD | *Pinus sylvestris* |
| UO | *Trifolium repens* |
| UO | *Taraxacum officinale* |
| UO | *Ranunculus repens* |
| UO | *Rumex acetosa* |
| UO | *Bellis perennis* |
| UO | *Stellaria media* |
| UO | *Plantago lanceolata* |
| WA | *Cirsium arvense* |
| WA | *Rumex acetosa* |
| WA | *Urtica dioica* |
| WA | *Plantago lanceolata* |
| WA | *Taraxacum officinale* |
| WA | *Heracleum phondylium* |
| WA | *Cerastium fontanum* |
| WA | *Trifloum repens* |
| WA | *Ranunculus repens* |
| WA | *Jacobaea vulgaris* |
| WA | *Quercus robur* |
| WA | *Quercus robur* |
| WA | *Picea abies* |
| WY | *Rubus fruticosus* |
| WY | *Corylus avellana* |
| WY | *Lonicera periclymenum* |
| WY | *Sambucus nigra* |
| WY | *Hedera helix* |
| WY | *Urtica dioica* |
| WY | *Glechoma hederacea* |
| WY | *Nepeta cataria* |
| WY | *Trifolium repens* |
| WY | *Taraxacum officinale* |
| WY | *Ranunculus repens* |
| WY | *Plantago major* |
| WY | *Rumex acetosa* |
| WY | *Heracleum sphondylium* |
| WY | *Petroselinum crispum* |
| WY | *Frangula alnus* |
| WY | *Polygonatum multiflorum* |
| WY | *Rosa canina* |
| WY | *Quercus robur* |
| WY | *Picea abies* |

**Table S 4: Unique compounds present per location.**

| Name | Location Name | Compound class |
|---|---|---|
| 1-decene | **OxBG Outdoors** | **Aliphatics** |
| 2,5-dimethylundecane | **OxBG Outdoors** | **Aliphatics** |

| | | |
|---|---|---|
| 3,5-dimethyloctane | **OxBG Outdoors** | **Aliphatics** |
| 4-methyltridecane | **OxBG Outdoors** | **Aliphatics** |
| Oxolane | **OxBG Outdoors** | **Aliphatics** |
| 1,4-diethylbenzene | **OxBG Outdoors** | **Aromatics** |
| 1h-indene | **OxBG Outdoors** | **Aromatics** |
| 2,5-dimethylfuran | **OxBG Outdoors** | **Aromatics** |
| Benzonitrile | **OxBG Outdoors** | **Aromatics** |
| Cumene | **OxBG Outdoors** | **Aromatics** |
| M-cymene | **OxBG Outdoors** | **Aromatics** |
| P-isopropenylmethylbenzene | **OxBG Outdoors** | **Aromatics** |
| Styrene | **OxBG Outdoors** | **Aromatics** |
| Tetralin | **OxBG Outdoors** | **Aromatics** |
| 2-thujene | **OxBG Outdoors** | **Monoterpenes** |
| Bornane | **OxBG Outdoors** | **Monoterpenes** |
| Prehnitene | **OxBG Outdoors** | **Monoterpenes** |
| Verbenone | **OxBG Outdoors** | **Monoterpenes** |
| Bromodichloromethane | **OxBG Outdoors** | **Others (Esters, N, S, Halo)** |
| Tolylacetate | **OxBG Outdoors** | **Others (Esters, N, S, Halo)** |
| Trichloroethene | **OxBG Outdoors** | **Others (Esters, N, S, Halo)** |
| Farnesane | **OxBG Outdoors** | **Sesquiterpenes** |
| 2,4-dimethyl-1-heptene | **OxBG Rainforest Glasshouse** | **Aliphatics** |
| 2,6,11-trimethyldodecane | **OxBG Rainforest Glasshouse** | **Aliphatics** |
| 3-hydroxy-3-methylbut-1-ene | **OxBG Rainforest Glasshouse** | **Aliphatics** |
| 3-pentanone, 2-methyl - | **OxBG Rainforest Glasshouse** | **Aliphatics** |
| Diacetone alcohol | **OxBG Rainforest Glasshouse** | **Aliphatics** |
| Hexane-2,3-dione | **OxBG Rainforest Glasshouse** | **Aliphatics** |
| Isoamylol | **OxBG Rainforest Glasshouse** | **Aliphatics** |
| Methyl salicylate | **OxBG Rainforest Glasshouse** | **Aromatics** |
| Musk gx | **OxBG Rainforest Glasshouse** | **Aromatics** |
| Alpha-terpinene | **OxBG Rainforest Glasshouse** | **Monoterpenes** |
| Beta-myrcene | **OxBG Rainforest Glasshouse** | **Monoterpenes** |
| Beta-ocimene | **OxBG Rainforest Glasshouse** | **Monoterpenes** |
| Beta-phellandrene | **OxBG Rainforest Glasshouse** | **Monoterpenes** |
| Gamma-terpinene | **OxBG Rainforest Glasshouse** | **Monoterpenes** |
| Sabinene | **OxBG Rainforest Glasshouse** | **Monoterpenes** |
| Isoamyl acetate | **OxBG Rainforest Glasshouse** | **Others (Esters, N, S, Halo)** |
| Propyl propionate | **OxBG Rainforest Glasshouse** | **Others (Esters, N, S, Halo)** |
| Copaene | **OxBG Rainforest Glasshouse** | **Sesquiterpenes** |
| Delta-cadinene | **OxBG Rainforest Glasshouse** | **Sesquiterpenes** |
| 4-terpineol | **OxBG Rainforest Glasshouse** | **Terpenoids** |
| Dihydromyrcenol | **OxBG Rainforest Glasshouse** | **Terpenoids** |
| Linalyl acetate | **OxBG Rainforest Glasshouse** | **Terpenoids** |
| Pinocarvone | **OxBG Rainforest Glasshouse** | **Terpenoids** |
| Sulcatone | **OxBG Rainforest Glasshouse** | **Terpenoids** |
| 2-methylbutan-1-ol | **University Parks Dense** | **Aliphatics** |
| 1h-1,2,4-triazole | **University Parks Open** | **Aromatics** |
| Butan-1-ol | **Warneford** | **Aliphatics** |
| Icosane | **Warneford** | **Aliphatics** |
| Tridecanal | **Warneford** | **Aliphatics** |
| 2-phenylpropan-2-ol | **Warneford** | **Aromatics** |
| 1-tetradecene | **Wytham** | **Aliphatics** |

| | | |
|---|---|---|
| Alpha-thujone | **Wytham** | **Terpenoids** |
